# The association between alcohol consumption and colorectal carcinogenesis is partially mediated by the gut microbiome

**DOI:** 10.1101/2024.10.17.24315656

**Authors:** Ane Sørlie Kværner, Einar Birkeland, Ekaterina Avershina, Edoardo Botteri, Cecilie Bucher-Johannessen, Markus Dines Knudsen, Anette Hjartåker, Christian M. Page, Johannes R. Hov, Mingyang Song, Kristin Ranheim Randel, Geir Hoff, Trine B. Rounge, Paula Berstad

**Author notes:** Corresponding Authors: Ane Sørlie Kværner, Mailing address: Cancer Registry of Norway, Norwegian Institute of Public Health, Postboks 5313 Majorstuen, 0304 Oslo, Norway, E-mail address, Paula Berstad, Mailing address: Cancer Registry of Norway, Norwegian Institute of Public Health, Postboks 5313 Majorstuen, 0304 Oslo, Norway.

## Abstract

**Background:** Alcohol consumption is one of the major risk factors of colorectal cancer (CRC). However, the mechanisms underlying this relationship are not fully understood, particularly the potential role of gut microbes.

**Objective:** To study associations of alcohol intake with the gut microbiome and colorectal lesions among CRC screening participants. Of particular interest was the potential role of gut microbes in mediating the association between alcohol intake and colorectal lesions.

**Methods:** Participants included fecal immunochemical test-positive women and men enrolled in the CRCbiome study, aged 55-77 years at inclusion. Intake of alcohol was assessed using a validated, semi-quantitative food frequency questionnaire. Integrating with shotgun metagenome based taxonomic and functional profiles, we studied associations with screen-detected colorectal lesions. The potential role of alcohol-associated gut microbes in mediating the association between alcohol intake and colorectal lesions was examined using causal mediation analysis.

**Results:** Of 1,468 participants with dietary data, 414 were diagnosed with advanced lesions (advanced adenoma, advanced serrated lesions or CRC). Alcohol intake was positively associated with advanced lesions in a dose-dependent manner (*p_trend_* = 0.008), with odds ratio of 1.09 (95% confidence interval, 1.00, 1.19) per 10 g/day increase. Compared to non-consumers, those consuming alcohol were characterized by a distinct microbial profile, manifested as modest, but consistent, shifts in α- and β-diversity, and differentially abundant bacteria (Log2 fold change (Log2FC) >0: *B. finegoldii* and *L. asaccharolyticus*; Log2FC <0: *S. mutans, B. dentinum*, *C. symbiosum* and *E. boltae*). A causal mediation analysis showed that 12% of the association between alcohol intake and advanced lesions was mediated by alcohol-associated gut bacteria.

**Conclusions:** Alcohol consumption was associated with a distinct microbial profile, which partly explained the association between alcohol intake and advanced colorectal lesions.

**Trial Registration:** The BCSN is registered at clinicaltrials.gov (National clinical trial (NCT) no. 01538550).

## Introduction

Alcohol increases the risk of cancer at multiple sites, in particular those of the gastrointestinal system^1–3^. According to recent global estimates, approximately 1 in 20 cancers in 2020 could be attributed to alcohol consumption^4^. Considering the rise in adult per capita consumption seen worldwide, especially in developing countries, these numbers are likely to increase in the years to come^5^. The changing patterns of alcohol consumption among women represent a particular cause of concern^5^.

Alcohol (ethanol) is efficiently absorbed by diffusion in the upper gastrointestinal tract, mainly in the stomach and small intestine, before entering the liver via the portal vein^6^. The predominant pathway for alcohol metabolism involves the enzymes alcohol dehydrogenase (ADH) and acetaldehyde dehydrogenase (ALDH), converting ethanol to acetaldehyde and acetate, respectively^6^. Alcohol can also be metabolized to acetaldehyde through the cytochrome P450 2E1 (CYP2E1) pathway^7^. Although the liver is the primary site for alcohol metabolism, some alcohol is also catabolized in the gastrointestinal tract, either by mucosal cells lining the gut or bacteria expressing enzymes involved in alcohol metabolism^7^.

Alcohol may promote cancer either directly or indirectly through its metabolites (acetaldehyde and acetate) and/or enzymes involved in alcohol metabolism^7^. Their potential carcinogenic effects are multiple, encompassing genomic, biochemical, inflammatory and immune-modulatory mechanisms, among others^7^. More recently, the gut microbiome has emerged as a plausible pathway through which alcohol may promote cancer^8^. However, it remains unclear whether and how alcohol contributes to carcinogenesis through this microbial pathway.

To gain deeper insight into the potential role of gut bacteria in alcohol-associated carcinogenesis, colorectal cancer (CRC) represents a particularly relevant malignancy. Not only are incidence rates highly connected to alcohol consumption (with approximately one in ten new cancer cases attributable to alcohol consumption^4^), but substantial data also support gut microbes as key players in the development of the disease^9^.

In the current study, we combined data on alcohol consumption with metagenome based taxonomic and functional profiles from participants in a large bowel cancer screening trial in Norway to shed light on this interplay. Specifically, our objectives included: I) investigating associations between alcohol consumption and the occurrence of screening-detected colorectal lesions, II) identifying microbial features linked to alcohol consumption, and III) evaluating whether potential associations between alcohol consumption and colorectal lesions are mediated by the microbiome.

## Subjects and Methods

### Bowel Cancer Screening in Norway (BCSN) and the CRCbiome study

The CRCbiome study is nested within the Bowel Cancer Screening in Norway (BCSN) study^10,11^, a pilot for the Norwegian national screening program. BCSN is a randomized trial comparing once-only sigmoidoscopy with four rounds of biennial fecal immunochemical testing (FIT). The BCSN trial was initiated in 2012 and has invited 139,291 women and men to participate. Of these, 70,096 were included in the FIT arm. FIT-positive participants (≥15 µg hemoglobin/g feces) were referred for work-up colonoscopy.

The CRCbiome study was initiated in 2017 and has during its four-year recruitment period invited 2,700 participants (starting from the second FIT round). The long-term goal of CRCbiome is to develop a microbiome-based biomarker to improve the current FIT based testing^11^. FIT-positive participants were invited to CRCbiome in the interval between being informed about their FIT screening result and attending colonoscopy. Besides the invitation letter, participants received two questionnaires to be completed prior to the colonoscopy examination: a food frequency questionnaire (FFQ) and a lifestyle and demographics questionnaire. Returning at least one of these questionnaires was regarded as consent to the study, being fulfilled by 1,640 (61%) participants. The age range at enrollment was 55-77 years.

Both the BCSN and the CRCbiome study have been approved by the Regional Committee for Medical Research Ethics in South-East Norway (Approval no.: 2011/1272 and 63148, respectively). The BCSN is also registered at clinicaltrials.gov (Clinical Trial (NCT) no.: 01538550).

### Study sample

The current study included participants from the CRCbiome study with available dietary information (n=1,616). After excluding participants who had withdrawn from the study after inclusion (n=15), not attended colonoscopy (n=39), had a poor quality FFQ (n=21) or reported too low (<600 and <800 kcal/day for women and men, respectively, n=9) or too high (>3,500 and >4,200 kcal/day for women and men, respectively, n=46) energy intake^12^, a final number of 1,486 participants were eligible for the study, including 947 individuals with available gut metagenome data (see flowchart, Figure 1).

**Figure 1.**
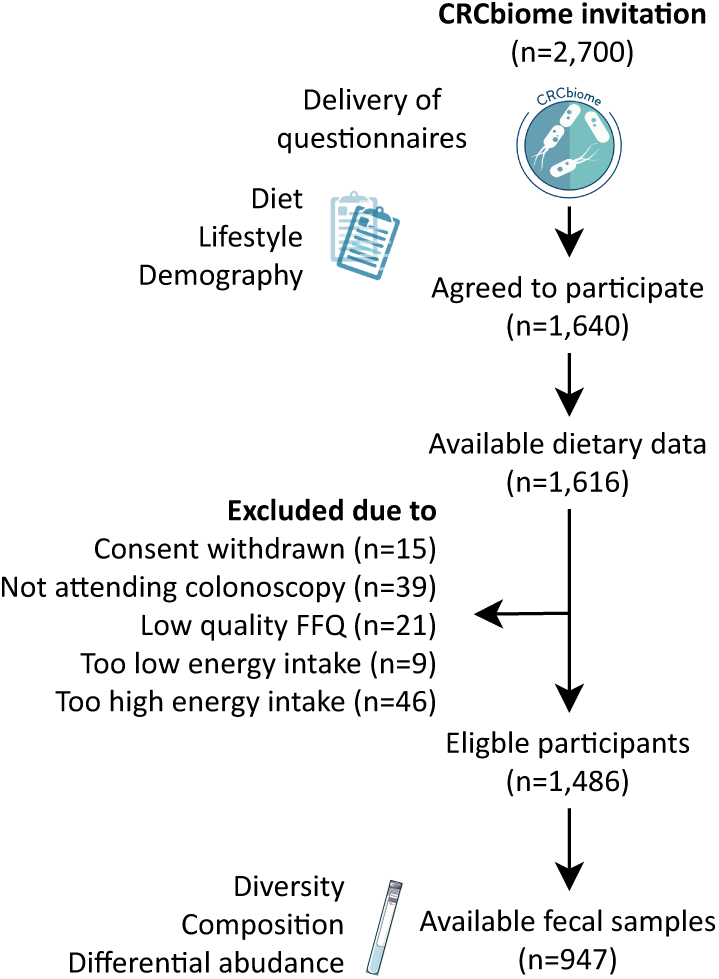
Flowchart of study participants.

### Assessment of alcohol intake

Information on dietary intake, including alcohol, was obtained using a self-administered semiquantitative, 14-page FFQ, designed to capture the habitual diet during the preceding year. The questionnaire is a modified version of an FFQ developed by the Department of Nutrition, University of Oslo^13–19^, which has been validated for a variety of nutrients^13,15,18,19^ and food groups^15–19^, including alcohol intake^13–15^. The questionnaire covers a total of 256 food and beverage items, of which eight concern alcoholic beverages (with one additional item covering non-alcoholic drinks). For each beverage type, participants were asked to record frequency of consumption, ranging from never/seldom to several times a week, and amount, typically as standardized alcoholic units. Daily alcohol intake was calculated using the dietary calculation system KBS (short for “**K**ost**b**eregnings**s**ystem”), developed at the Department of Nutrition, University of Oslo. The most recent database at the time, AE-18, was used. AE-18 is an extended version of the official Norwegian Food Composition Table, version 2018^20^. Alcohol intake was quantified both as ethanol (g/day and energy percentage (E%)) and by alcoholic beverage type (g/day). The alcoholic beverages were categorized as follows (with the standardized unit used in calculations given in brackets): ‘Wine’, including both red and white wine (15 cl); ‘beer’, comprising regular and light beer (33 cl); ‘spirits’, encompassing spirits, mulled wines like port and cherry, and liqueurs (4 cl); and ‘drinks’, which included cocktails, alcoholic cider, and alcohol-containing soft drinks (20 cl). Additionally, the ‘non-alcoholic drinks’ category, consisting of non-alcoholic beers (33 cl), was evaluated for comparison purposes. One alcoholic unit was defined as 12 g^21^.

Prior to analyses, all questionnaires were reviewed and evaluated by trained personnel according to a standardized framework for quality control assessment developed by the study group^11^.

### Outcome assessment

Outcome data were obtained from the BCSN database, containing detailed clinicopathological information on all colorectal lesions detected at work-up colonoscopy. The information was recorded by the responsible endoscopist using a structured reporting system. Based on the most severe finding at colonoscopy, participants were categorized into the following three diagnostic groups: advanced lesions, comprising CRC (any adenocarcinoma of the colon or rectum), advanced adenomas (any adenoma with villous histology, high-grade dysplasia or diameter ≥10 mm) and advanced serrated lesions (any serrated lesion with size ≥10 mm or dysplasia); non-advanced adenomas; and controls, i.e., no CRC, adenoma nor advanced serrated lesions detected.

### Sample collection, library generation and shotgun metagenome sequencing

Protocols for sample collection, library generation and shotgun metagenome sequencing have been described in detail elsewhere^11^. In brief, DNA was extracted from 500 µl aliquots, derived from left-over buffer of FIT samples, using the QIAsymphony DSP Virus/Pathogen Midikit (Qiagen, Hilden, Germany), with an offboard lysis protocol based on bead-beating. Purified DNA was further eluted in 60 µl AVE buffer (Qiagen, Hilden, Germany). Following extraction, DNA concentration was measured on Qubit (Thermo Fisher Scientific, MA, USA). For samples with a DNA concentration <1.5 ng/µl, DNA was extracted from a second aliquot. FIT samples with a concentration of 0.7 ng/µl or more were considered eligible for shotgun metagenomic library preparation, whereas for samples with DNA extracted from multiple aliquots, the one with the highest DNA concentration was used.

Sequencing libraries were generated according to the Nextera DNA Flex Library Prep Reference Guide, except scaling down the reaction volumes to one fourth of the reference. Library pools of 240 samples were combined and size selected to a fragment size of 650 – 900 bp. Sequencing was performed on the Illumina NovaSeq system (Illumina Inc., CA, USA) using S4 flow cells with lane divider, with each pool sequenced on a single lane resulting in paired end 2 x 151 bp reads.

### Determination of taxonomic and functional profiles

Sequencing reads were processed for removal of adapters and low-quality bases using trimmomatic (v0.36)^22^ with the following trimming options: leading 20, trailing 20, minlength 50. Reads mapping to the human genome (hg38) and PhiX were removed using Bowtie2 (v2.3.5.1)^23^. Read-based taxonomy and gene content was assessed using MetaPhlAn3 (v3.0.7) and HumanN3 (v3.0.0)^24^, respectively, with the mpa v30 ChocoPhlAn 201901 pangenome database, using the UniRef90 database to assign gene families to Metacyc pathways. Read-based taxonomic abundance was evaluated at the species level. Pathway abundance was scaled by the number of quality-controlled reads per million.

### Assessment of covariates

Information on covariates was obtained using a self-administered, four-page questionnaire on lifestyle and demographic data, which has been described in detail previously^11^. The questions relevant to the current study concerned demographic factors (national affiliation, education, occupation and marital status), clinical factors (family history of CRC and diagnosis of chronic bowel disorders) and lifestyle factors (smoking and snus habits and physical activity level).

Smokers and snusers were defined as self-reported regular or occasional users or those having quit consumption within the last ten years. Total amount of moderate to vigorous physical activity (minutes/week) was calculated by summing the time spent in moderate and vigorous activity, the latter weighted by a factor of two to best match recent guidelines^25–27^. Body mass index (BMI) was calculated based on self-reported weight (kg) and height (cm) obtained from the FFQ.

### Statistics

Descriptive statistics are presented as median and interquartile range (Q1, Q3) for continuous variables and numbers and percentages for categorical variables.

Pairwise correlations between continuous measures were computed using Spearman’s correlation coefficients (*r*).

To study the association between alcohol intake and colorectal lesions, multinomial logistic regression analysis was conducted. Colonoscopy findings were categorized into advanced lesions, non-advanced adenomas and controls, according to the outcome definition given above. Alcohol (as ethanol in g/day) was categorized by consumption level (0 g/day, >0-10 g/day, ≥10- 20 g/day and ≥20 g/day) and by adherence to national guidelines (full adherence: 0 g/day, partial adherence: <10 and 20 g/day for women and men, respectively, and non-adherence: ≥10 and 20 g/day for women and men, respectively^21,25,28^). Linear (per 10 g/increase/day) and exponential (per 2-fold increase) consumption was evaluated, the latter based on a log2-transformation of the continuous alcohol variable in g/day plus 1 g/day, to enable inclusion of non-consumers in the analysis. For the alcohol subtypes, participants were categorized as consumers/non-consumers.

The selection of covariates was based on *a priori* knowledge on the relationship between alcohol intake and colorectal lesions^29–31^, with all multinomial logistic regression analyses being adjusted for the following covariates: age (continuous), sex (women, men), national affiliation (Norwegian, non-Norwegian, missing), screening center (center 1, center 2), education level (primary school, high school, college/university, missing), family history of CRC (yes, no, unknown/missing), smoking status (non-smoker, smoker, missing), BMI (continuous, with missing set to median) and level of physical activity (continuous, with missing set to median).

Additional adjustment for potential dietary risk or protective factors, such as red and processed meat, whole grain, and dairy products – as well as alternative ways of categorizing covariates, including a more refined smoking variable, were also evaluated, but not included in the final model as they did not influence the interpretation of results.

To study potential differential influence of alcohol intake on colorectal lesions by sex, separate analyses were conducted in women and men. Potential interactions were examined using the Wald test. Subgroup analyses were also conducted by precursor lesion subtype (advanced adenoma or advanced serrated lesion) and location (advanced proximal or advanced distal lesion).

As sensitivity analyses, the main association analyses were run with alcohol intake calculated as E% rather than g/day, use of a multiple imputation approach for handling of missing data, or the exclusion of participants without metagenome data (n=539). The potential influence of leaving out participants with self-reported bowel disorders (n=216) was also evaluated.

To identify microbial features linked to higher alcohol intake, associations of alcohol with the following three measures were examined: α-diversity by means of the Shannon and inverse Simpson indices, β-diversity based on the Bray-Curtis dissimilarity metric, and bacterial species and pathway abundance. Associations with α-diversity were examined using linear regression models with the diversity indices as the dependent variable. To improve normality and ease interpretation, diversity indices were log-transformed prior to analysis. β-diversity was evaluated by principal coordinate analysis (PCoA) and permutational multivariate analysis of variance (PERMANOVA) with 999 permutations. Effect sizes were determined by calculating the partial omega-squared (Ω^2^) values. Differential abundance analyses were performed using microbiome multivariable associations with linear models (MaAsLin) 2^32^ with the following settings: Min. abundance: 0.0; min. prevalence: 0.1; normalization: none and total sum scaling (TSS) for bacteria and pathways, respectively; transformation: log2-transformation with a pseudo-count of half the minimum value; and analyzing method set to a linear model. Benjamini-Hochberg corrected p-values were used as basis for interpretation of results. All analyses on microbial data were conducted in the study group as a whole and stratified by sex. The same set of covariates were adjusted for as in the association analyses with lesions as outcome variable, but with the addition of sequencing depth as a continuous variable. Other combinations of covariates were also evaluated (i.e. the addition of dietary risk or protective factors (as elaborated on above), presence of bowel disorders, as an indicator of bowel movement pattern, and antibiotic usage), but not included in subsequent analyses as they only marginally altered the results.

To evaluate whether the gut microbiome mediated the association between alcohol intake and advanced lesions, a causal mediation analysis was applied. An alcohol-associated microbial score was developed as an indicator of the effect of alcohol consumption on the gut microbiome. This score was constructed based on the output of adjusted differential abundance analyses, following the formula proposed by Gevers, *et al*.^33^:

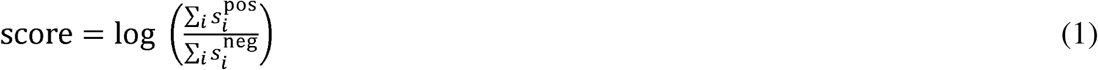

where 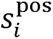 and 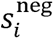 were the abundance of species positively and negatively associated with any level of alcohol consumption, respectively. Zeros in either the numerator or denominator were avoided by substituting zeros with a pseudo-count of half the minimum value of the numerator or denominator across the dataset, respectively. To minimize bias in the estimated effects, a five-fold cross-validation approach was employed for generating the microbial score. Specifically, the dataset was divided into five non-overlapping subsets, where for each subset, a microbial score was calculated using the significantly associated microbial species identified from a fully adjusted differential abundance analysis conducted on the remaining four subsets. The significance threshold for this analysis was set to a Benjamini-Hochberg-adjusted p-value of 0.1.

In the causal mediation analysis, 2-fold increases in alcohol intake were treated as the independent variable, advanced lesions as the dependent variable and the alcohol-associated microbial score as the potential mediator. Non-advanced adenomas were grouped with colonoscopy-negatives for these analyses. The mediation analysis was adjusted for the same set of covariates as in the multinomial logistic regression models, as well as the cross-validation partition identity. We used the R package mediation^34^ and the function ‘*mediate*’ based on the following two models: 1) a multivariate linear regression model examining the association between the exposure (E) and the mediator (M), and 2) a multivariate generalized linear model examining the association between the mediator (M) and the outcome (O), controlling for the exposure (E). We report the point estimates along with non-parametric bootstrap confidence intervals using the percentile method. Uncertainty estimates were calculated using 1000 simulations. We also employed the medflex R package^35^ as an alternative approach to mediation analysis. In this analysis we used a natural effects model with imputation of nested counterfactuals, ensuring that mediation effects were not dependent on covariate levels.

Statistical analyses were performed using R, version 4.1.0 (The R Foundation for Statistical Computing, Vienna, Austria). In addition to packages included in tidyverse (version 1.3.1), main packages, with version number in parentheses, included skimr (2.1.5), corrplot (0.88), nnet (7.3- 16), VGAM (1.1-5), mice (3.13.0), vegan (2.5-7), micEco (0.9.15), Maaslin2 (1.12.0), mediation (4.5.0) and medflex (0.6-7).

## Results

### Study population

**Table 1** shows the key characteristics of the study population overall and by alcohol intake. The median (Q1, Q3) age of participants was 67 (62, 72) years, with a slightly higher representation of men (56%). Compared to non-consumers (13% of participants), those consuming alcohol were more likely to be male and be affiliated to the screening center localized in Bærum (center 2).

**Table 1.** Key characteristics of the study population overall and by alcohol intake (n=1,486)^1^.

| Variables | n | Level of alcohol intake |  |  |  |  |
| --- | --- | --- | --- | --- | --- | --- |
|  |  | Overall<br>(n=1,486) | 0 g/day<br>(n=187) | >0-10 g/day<br>(n=592) | ≥10-20 g/day<br>(n=361) | ≥20 g/day<br>(n=346) |
| Age, years | 1,486 | 67 (62, 72) | 67 (62, 73) | 67 (62, 72) | 66 (62, 72) | 68 (62, 72) |
| Male sex, n (%) | 1,486 | 826 (55.6) | 70 (37.4) | 281 (47.5) | 212 (58.7) | 263 (76.0) |
| Screening center, n (%) | 1,486 |  |  |  |  |  |
| Center 1 (Moss) |  | 774 (52.1) | 135 (72.2) | 323 (54.6) | 166 (46.0) | 150 (43.4) |
| Center 2 (Bærum) |  | 712 (47.9) | 52 (27.8) | 269 (45.4) | 195 (54.0) | 196 (56.6) |
| National affiliation, n (%) | 1,427 |  |  |  |  |  |
| Norwegian |  | 1345 (94.3) | 160 (89.4) | 540 (94.7) | 333 (96.0) | 312 (94.3) |
| Non-Norwegian |  | 82 (5.7) | 19 (10.6) | 30 (5.3) | 14 (4.0) | 19 (5.7) |
| Family history of CRC, n (%) | 1,347 |  |  |  |  |  |
| No |  | 1,092 (81.1) | 133 (83.1) | 442 (81.1) | 272 (83.2) | 245 (77.8) |
| Yes |  | 255 (18.9) | 27 (16.9) | 103 (18.9) | 55 (16.8) | 70 (22.2) |
| Marital status, n (%) | 1,465 |  |  |  |  |  |
| Married/cohabiting |  | 1,171 (79.9) | 123 (67.6) | 460 (79.0) | 308 (85.8) | 280 (81.9) |
| Not married/non-cohabiting |  | 294 (20.1) | 59 (32.4) | 122 (21.0) | 51 (14.2) | 62 (18.1) |
| Education, n (%) | 1,462 |  |  |  |  |  |
| Primary school |  | 251 (17.2) | 54 (29.8) | 102 (17.5) | 50 (14.0) | 45 (13.2) |
| High school |  | 580 (39.7) | 75 (41.4) | 243 (41.8) | 144 (40.2) | 118 (34.6) |
| University/college |  | 631 (43.2) | 52 (28.7) | 237 (40.7) | 164 (45.8) | 178 (52.2) |
| Working status, n (%) | 1,464 |  |  |  |  |  |
| Employed |  | 498 (34.0) | 40 (22.1) | 190 (32.6) | 140 (39.1) | 128 (37.4) |
| Retired/unemployed |  | 966 (66.0) | 141 (77.9) | 393 (67.4) | 218 (60.9) | 214 (62.6) |
| Bowel disorder, n(%) | 1,451 |  |  |  |  |  |
| No bowel disease |  | 1,235 (85.1) | 147 (82.6) | 482 (84.0) | 300 (83.8) | 306 (89.7) |
| IBS |  | 81 (5.6) | 10 (5.6) | 30 (5.2) | 31 (8.7) | 10 (2.9) |
| Celiac disease |  | 18 (1.2) | 3 (1.7) | 8 (1.4) | 4 (1.1) | 3 (0.9) |
| IBD |  | 24 (1.7) | 5 (2.8) | 11 (1.9) | 7 (2.0) | 1 (0.3) |
| Other |  | 93 (6.4) | 13 (7.3) | 43 (7.5) | 16 (4.5) | 21 (6.2) |
| Smoking status<sup>2</sup>, n (%) | 1,462 |  |  |  |  |  |
| Non-smoker |  | 1,082 (74.0) | 127 (70.2) | 438 (75.3) | 270 (75.4) | 247 (72.4) |
| Smoker |  | 380 (26.0) | 54 (29.8) | 144 (24.7) | 88 (24.6) | 94 (27.6) |
| Snus status<sup>3</sup>, n(%) | 1,405 |  |  |  |  |  |
| Non-snuser |  | 1,308 (93.1) | 171 (97.2) | 530 (95.3) | 318 (92.4) | 289 (87.8) |
| Snuser |  | 97 (6.9) | 5 (2.8) | 26 (4.7) | 26 (7.6) | 40 (12.2) |
| BMI, kg/m<sup>2</sup> | 1,480 | 26 (24, 29) | 27 (25, 30) | 26 (24, 29) | 26 (24, 29) | 27 (24, 29) |
| Physical activity, min/week | 1,466 | 135 (0, 300) | 45 (0, 195) | 135 (0, 300) | 180 (15, 390) | 180 (45, 352) |
| Questionnaires completed prior to colonoscopy, n(%) | 1,486 |  |  |  |  |  |
| No |  | 126 (8.5) | 15 (8.0) | 42 (7.1) | 30 (8.3) | 39 (11.3) |
| Yes |  | 1,360 (91.5) | 172 (92.0) | 550 (92.9) | 331 (91.7) | 307 (88.7) |

<sup>1</sup>Values are median (Q1, Q3) for continuous variables and n (%) for categorical variables.<sup>2,3</sup>To be defined as a smoker or snuser one had to be a regular or occasional user or having quit consumption within the last ten years.
Abbreviations: BMI; Body mass index, CRC; colorectal cancer, g; gram, n; number

Higher alcohol intakes were in general positively associated with markers of higher socioeconomic status such as being married or cohabiting, being employed, and holding a university or college degree. The high consumers were also more likely to use snus tobacco and reported having a higher level of physical activity than the non-consumers.

### Daily intake of alcohol

Daily intake of alcohol and different types of alcoholic beverages are presented in **Table 2**. The median (Q1, Q3) intake of alcohol (as ethanol in g/day) was 9 (2-19) g/day; 13 (4-25) g/day in men and 5 (1-13) g/day in women. Despite higher intakes in men, the proportion adhering to national guidelines was relatively similar between sexes (68% in men, 65% in women). In terms of beverage types, wine was the most frequently consumed by women (74%), whereas beer was the most frequently consumed by men (80%). There were fewer consumers of spirits (30%) and drinks (17%), particularly among women (17 and 13%, respectively). Alcohol (overall and by subtype) was only modestly correlated with energy intake (with Spearman’s *r*’s ranging from 0.06-0.24 in the study group as a whole).

**Table 2.** Alcohol consumption in the study population as a whole (n=1,486) and by sex (660 women, 826 men).

|  | Percentiles |  |  |  | Adherent to guidelines<sup>1</sup> | Zero consumers | Correlation with energy intake |
| --- | --- | --- | --- | --- | --- | --- | --- |
| | 25 | 50 | 75 | 100 | n (%) | n (%) | $r_s^2$ |
| **Overall** |  |  |  |  |  |  |  |
| Alcohol, g/day | 2.2 | 9.0 | 19 | 195 | 991 (67) | 187 (13) | 0.22** |
| Alcohol, E% | 0.7 | 2.9 | 6.0 | 45 | 1003 (68) | 187 (13) | -0.04 |
| Alcohol units<sup>3</sup>/day | 0.2 | 0.7 | 1.6 | 16 | 1083 (73) | 187 (13) | 0.22** |
| Alcohol subtypes, g/day |  |  |  |  |  |  |  |
| Beer | 0.0 | 33 | 140 | 3743 | - | 535 (36) | 0.24** |
| Wine | 0.0 | 36 | 108 | 837 | - | 417 (28) | 0.06* |
| Spirits | 0.0 | 0.0 | 1.2 | 223 | - | 1040 (70) | 0.11** |
| Drinks, cider, etc. | 0.0 | 0.0 | 0.0 | 768 | - | 1240 (83) | 0.10** |
| Non-alcoholic drinks, g/day | 0.0 | 0.0 | 14 | 2000 | - | 1100 (74) | 0.13** |
| **Men** |  |  |  |  |  |  |  |
| Alcohol, g/day | 3.8 | 13 | 25 | 195 | 563 (68) | 70 (8) | 0.17** |
| Alcohol, E% | 1.2 | 3.6 | 7.2 | 45 | 503 (61) | 70 (8) | -0.10* |
| Alcohol units<sup>3</sup>/day | 0.3 | 1.1 | 2.1 | 16 | 610 (74) | 70 (8) | 0.17** |
| Alcohol subtypes, g/day |  |  |  |  |  |  |  |
| Beer | 16 | 80 | 203 | 3743 | - | 167 (20) | 0.19** |
| Wine | 0 | 35 | 108 | 837 | - | 247 (30) | 0.04 |
| Spirits | 0 | 0 | 4,8 | 223 | - | 489 (59) | 0.07 |
| Drinks, cider, etc. | 0 | 0 | 0 | 768 | - | 667 (81) | 0.07* |
| Non-alcoholic drinks, g/day | 0 | 0 | 14 | 2000 | - | 591 (72) | 0.11* |
| **Women** |  |  |  |  |  |  |  |
| Alcohol, g/day | 1.1 | 5.4 | 13 | 87 | 428 (65) | 117 (18) | 0.13* |
| Alcohol, E% | 0.4 | 2.0 | 4.8 | 34 | 500 (76) | 117 (18) | -0.12* |
| Alcohol units<sup>3</sup>/day | 0.1 | 0.5 | 1.1 | 7.23 | 473 (72) | 117 (18) | 0.13* |
| Alcohol subtypes, g/day |  |  |  |  |  |  |  |
| Beer | 0.0 | 0.0 | 45 | 1280 | - | 368 (56) | 0.11* |
| Wine | 0.0 | 41 | 108 | 837 | - | 170 (26) | 0.09* |
| Spirits | 0.0 | 0.0 | 0.0 | 78 | - | 549 (83) | 0.04 |
| Drinks, cider, etc. | 0.0 | 0.0 | 0.0 | 288 | - | 571 (87) | 0.08 |
| Non-alcoholic drinks, g/day | 0 | 0 | 0 | 500 | - | 511 (77) | 0.19** |

<sup>1</sup><10 g/day for women and <20 g/day for men according to Nordic and national nutritional recommendations<sup>21,25</sup>; <5 energy percentage (E%) for both sexes according to Nordic and national nutritional recommendations<sup>21,25</sup>; <1 unit for women and <2 units for men according to national food-based dietary guidelines<sup>62</sup>.
<sup>2</sup>\*\*<0.001, \*<0.05.
<sup>3</sup>1 unit set to 12 grams in line with national guidelines<sup>21,25</sup>.
Abbreviations: E%; energy percentage, n; number, $r_s$ ; Spearman's correlation coefficient.

### Alcohol intake and colorectal lesions

Associations between alcohol intake (as ethanol in g/day) and colorectal lesions are shown in **Table 3**. Compared to non-consumption, all levels of alcohol intake, and in particular high levels, were positively associated with advanced lesions (*p_trend_* = 0.008). The probability of advanced lesions increased by 9% per 10 g increase/day and 14% per 2-fold increase/day. Partial and non-adherence to guidelines were also positively associated with advanced lesions relative to not consuming alcohol, with odds ratios (ORs) of 1.91 (95% confidence intervals (CI) 1.22, 2.99) and 1.97 (1.23, 3.17), respectively. Notable differences were observed between women and men, with associations being consistently stronger for women. As an example, relative to those not consuming alcohol, women in the highest consumption category (≥ 20 g/day) had an OR for advanced lesions of 4.81 (95% CI 2.18, 10.62) compared to 1.48 (0.74, 2.96) in men (*p_interaction_* = 0.040). No associations between alcohol intake and non-advanced adenomas were detected. Subgroup analyses by lesion subtype and location implied associations for both groups of precursor lesions, regardless of lesion location (**Supplementary Table 1**).

**Table 3.**
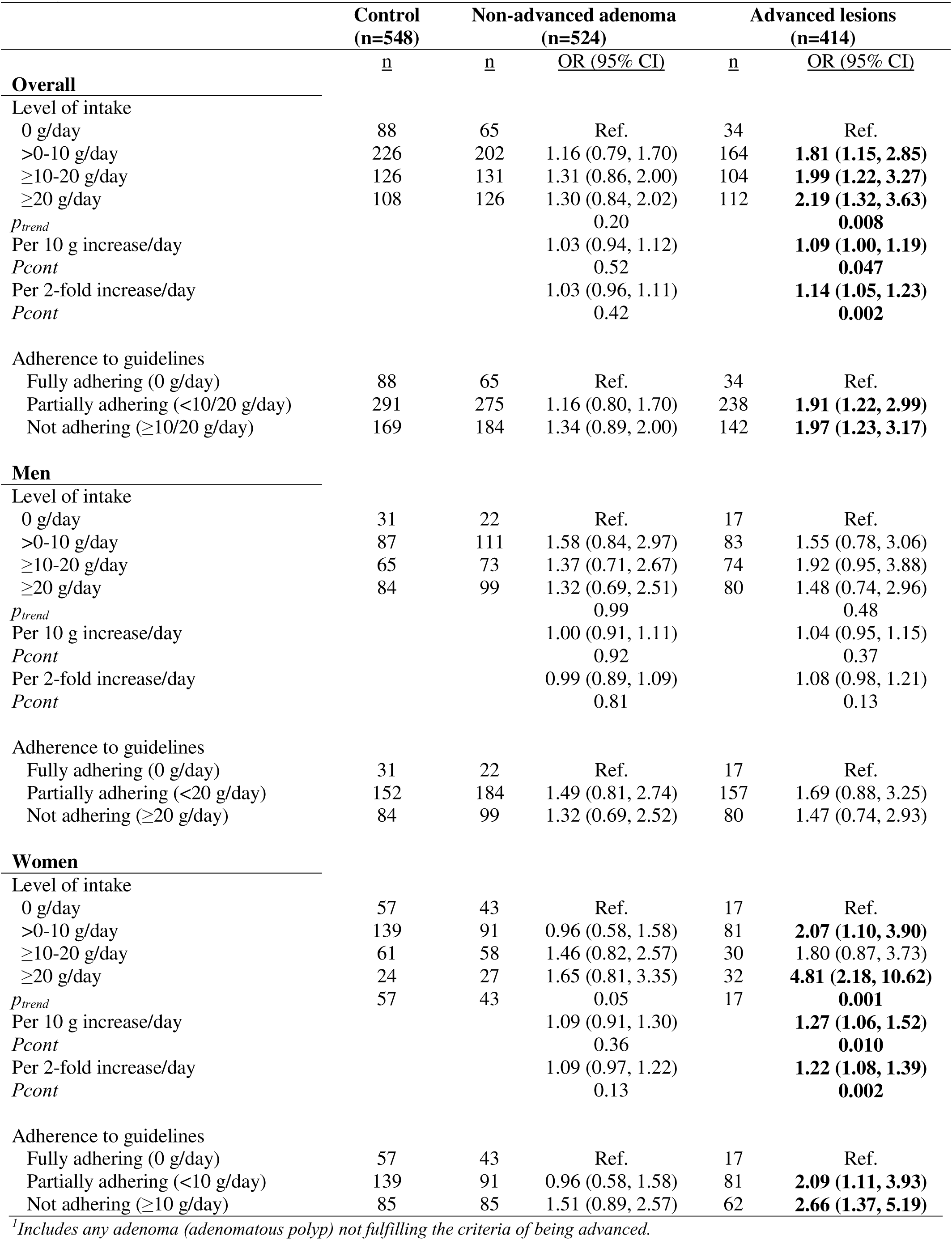
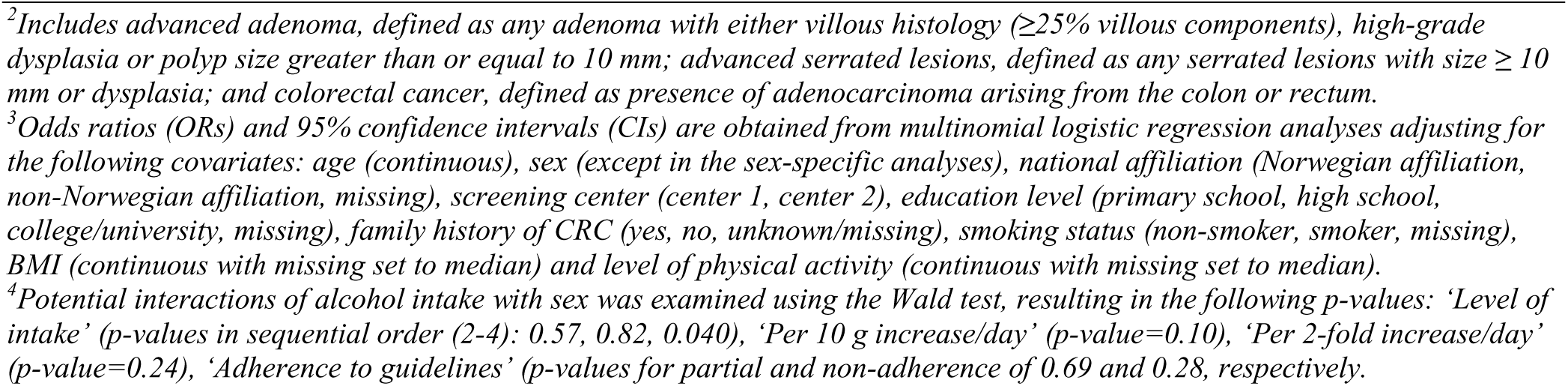
Odds ratios (ORs) and 95% confidence intervals (CIs) for presence of non-advanced adenoma^1^ and advanced lesions^2^ relative to controls by level of alcohol consumption in the study population as a whole (n=1,486) and by sex (660 women, 826 men)^3,4^.

Sensitivity analyses with alcohol intake as E%, use of a multiple imputation approach for handling of missing values and restricting the study population to those with metagenome data only (**Supplementary Tables 2-4**) produced similar results as in the main analyses. This was also the case for an analysis excluding participants with a self-reported bowel disorder (data not shown).

In terms of beverage types, positive associations with advanced lesions were seen for consumers of wine (OR 1.41; 95%CI 1.03, 1.92) and beer (OR 1.31; 0.97, 1.77; **Figure 2**). Stratifying the analyses by sex, the association for wine was stronger (and only statistically significant) in women, whereas the association for beer tended to be stronger for men (although not statistically significant). No associations with non-advanced adenomas were observed.

**Figure 2.**
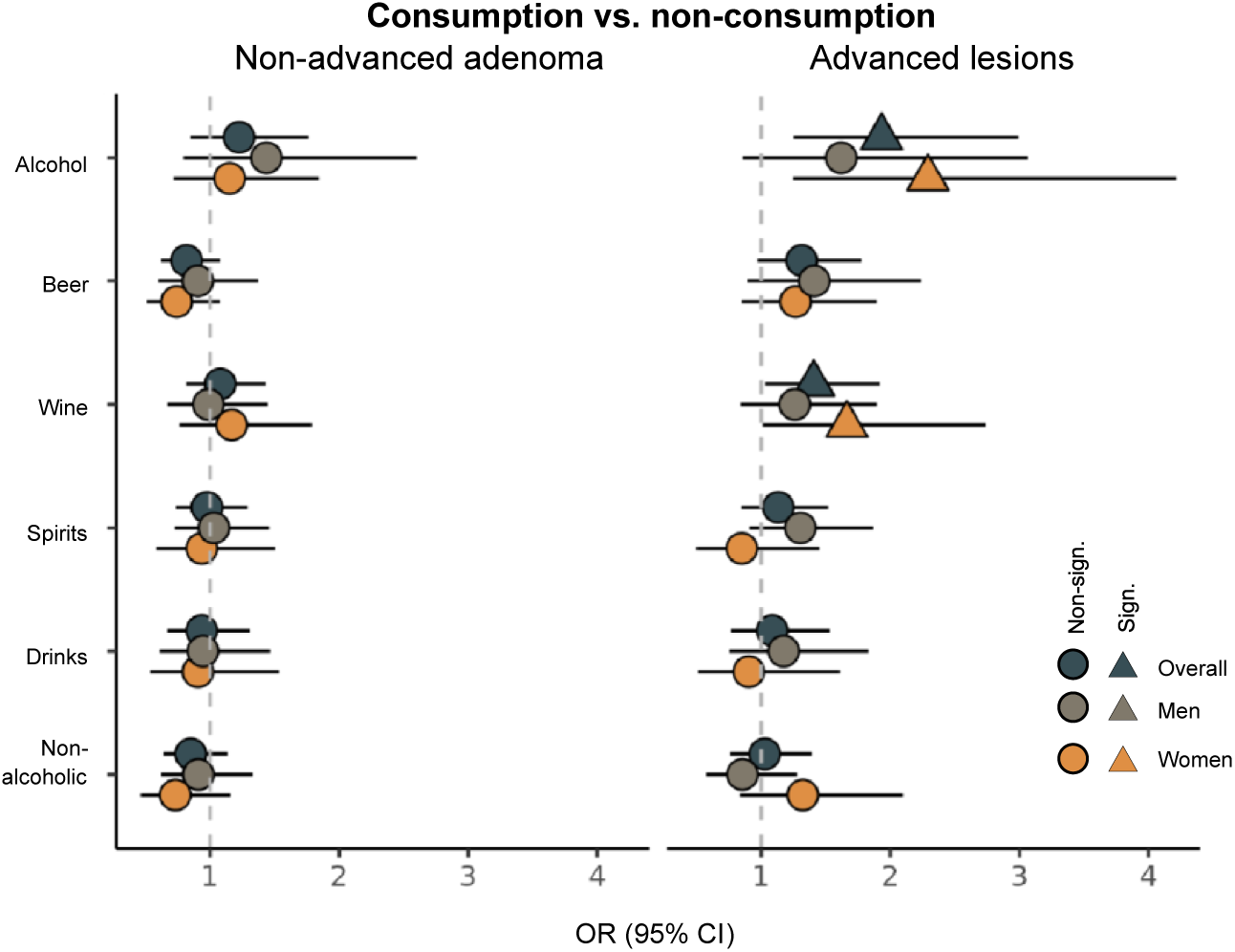
Odds ratios (ORs) and 95% confidence intervals (CIs) for presence of non-advanced adenoma and advanced lesions relative to controls for participants consuming *vs*. not consuming alcohol, as well as the different types of alcoholic beverages in the study overall (n=1,486) and by sex (660 women, 826 men).

### Alcohol intake and gut microbial features

Metagenome shotgun sequencing data derived from leftover buffer containing fecal matter collected in screening FIT cartridges was available for 947 participants (mean = 12 million paired end reads, sd = 3.8 million). Taxonomic classification resulted in identification of a total of 787 microbial species (mean = 88, sd = 15.5 per sample).

### α-diversity

Associations of alcohol intake with the α-diversity indices Shannon and Inverse Simpson are presented in **Figure 3a-c** and **Supplementary Table 5**. In general, there was a weak, but statistically significant positive association between alcohol intake (as ethanol in g/day) and both diversity indices. The shift in diversity became noticeable already at low intake levels. Compared to the non-consumers, those consuming alcohol had a 2.7 and 10.2% higher Shannon and Inverse Simpson index, respectively. No dose-response associations were observed. Stratification by sex showed the associations between total alcohol intake and α-diversity to be statistically significant in women only. Still, looking at alcoholic beverage subtypes, statistically significant positive associations were observed for consumers of beer (overall and in men), wine (women), spirits (overall and in women) and non-alcoholic drinks (women). As for ethanol in g/day, there were no clear dose-response relationships.

**Figure 3.**
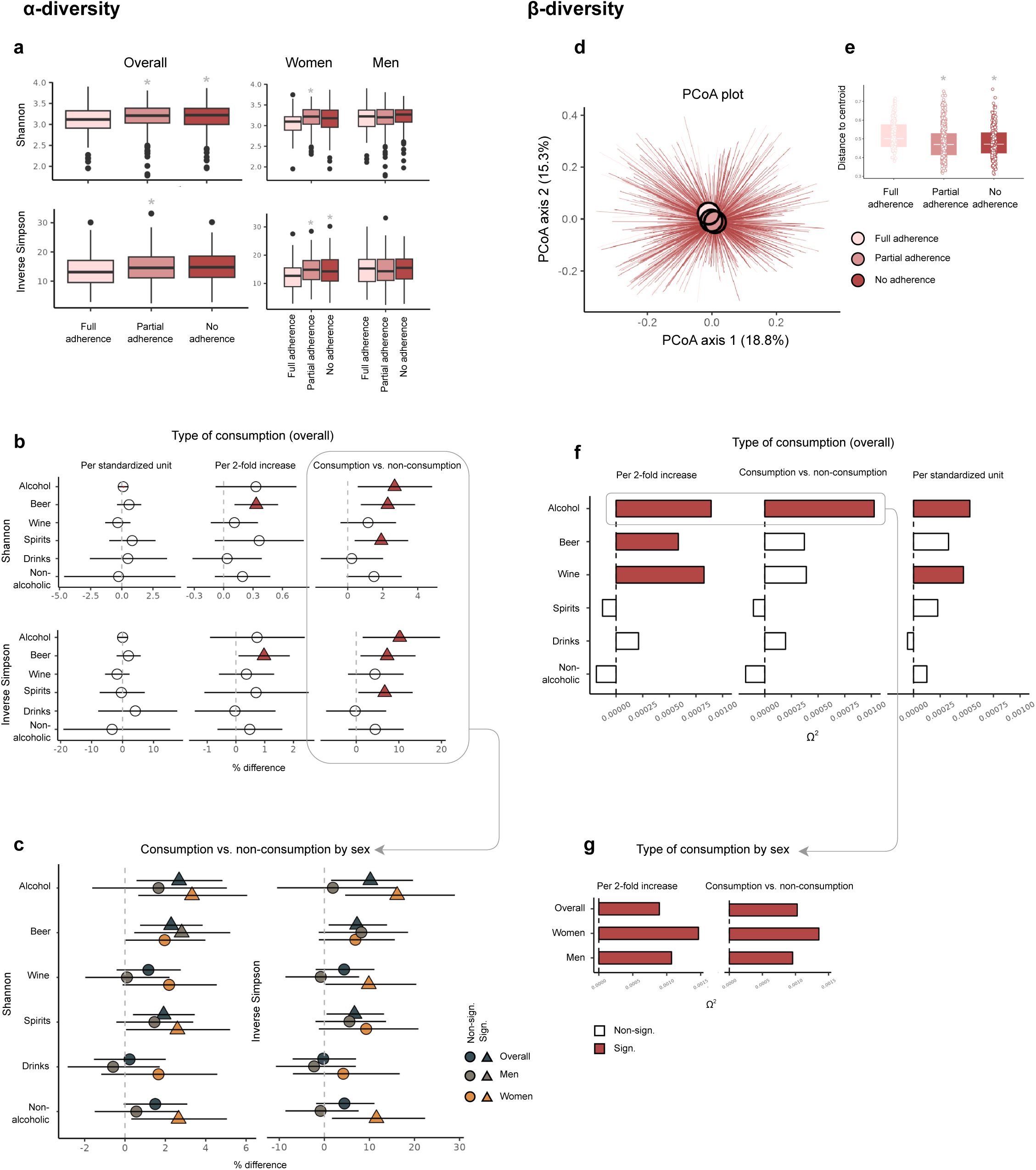
Associations of alcohol intake with the α-diversity indices Shannon and Inverse Simpson (a-c) and β- diversity based on the Bray-Curtis dissimilarity metric (d-g).

### β-diversity

Associations of alcohol intake with the Bray-Curtis β-diversity index are presented in Figure 3d- g and Supplementary Table 5. Irrespective of the approach to quantifying alcohol intake (as ethanol in g/day), the microbial composition differed by intake level (Ω values of 0.0005-0.0010, PERMANOVA-derived p-values <0.05, Supplementary Table 5). Consumers of alcohol displayed a small shift in microbial composition (Figure 3d), and were less heterogeneous (Figure 3e) than the non-consumers. There was, however, no difference in microbial composition by intake level (data not shown). Consuming alcohol was associated with microbial composition regardless of sex.

Consumption of wine, and to a lesser extent, beer, seemed to be related to microbial composition (Figure 3f).

#### Differentially abundant bacteria and pathways

Associations between alcohol intake and bacteria and pathway abundance are presented in **Figures 4a** and **5a**. In total, 6 bacteria (2 positively and 4 negatively) and 7 pathways (5 positively and 2 negatively) were statistically significantly associated with at least one of the alcohol consumption categories (Figures 4a and 5a). Of those predictive of the highest consumption category, 4 out of 5 bacteria (i.e. *L. asaccharolyticus*, *B. finegoldii*, *S. mutans* and *C. symbiosum*) and 3 out of 4 pathways (i.e. the tricarboxylic acid (TCA) cycle II, superpathway of sulfur oxidation and L-lysine biosynthesis II), remained statistically significant after mutual adjustment for the other bacteria and pathways, respectively (**Supplementary Figure 2a**).

**Figure 4.**
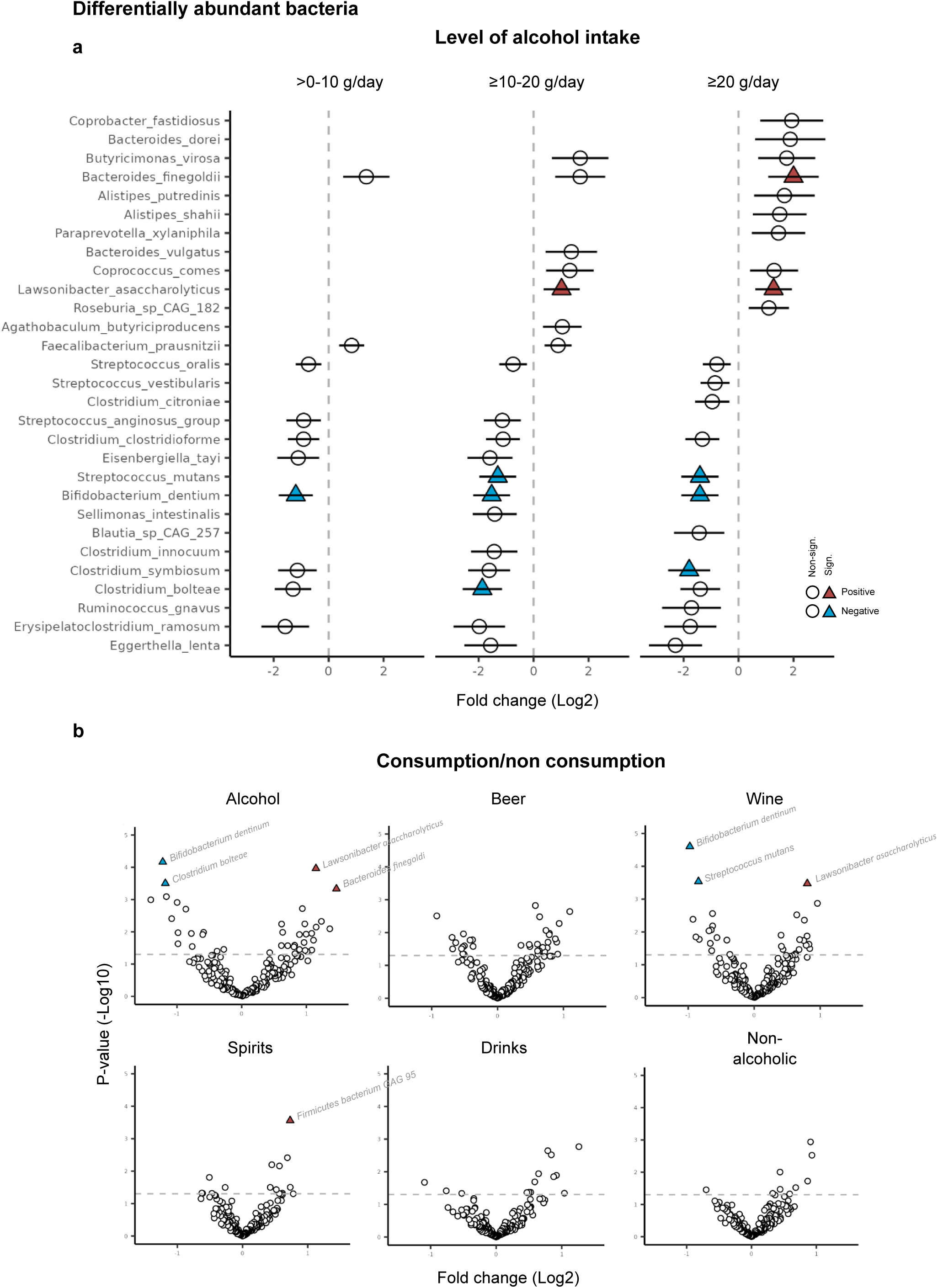
Differential abundance analyses of bacterial species by total alcohol intake and alcoholic beverage types.

**Figure 5.**
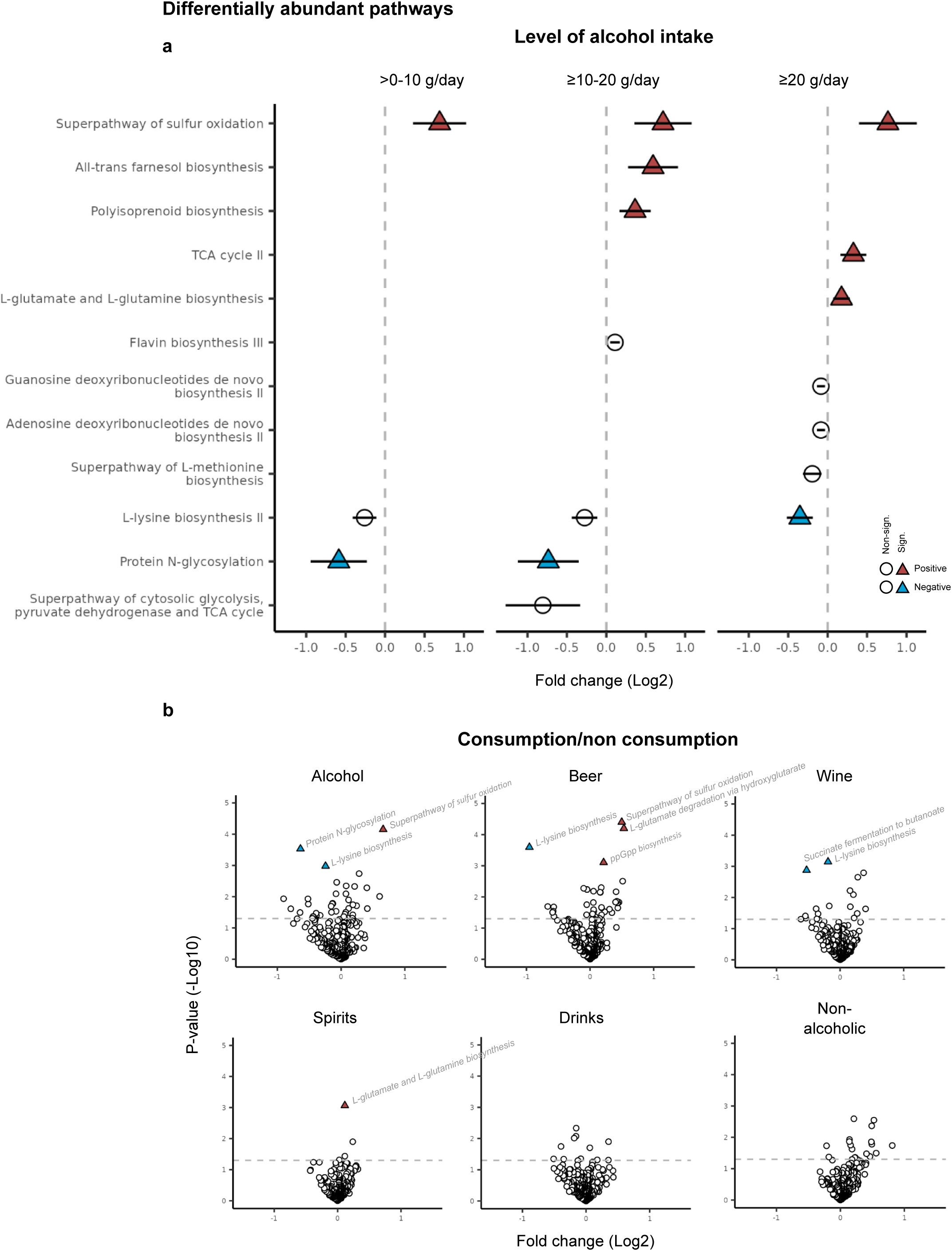
Differential abundance analyses of pathways by total alcohol intake and alcoholic beverage types.

Several of the identified bacteria and pathways were inter-correlated (Supplementary Figure 2b). None of the identified bacteria or pathways showed signs of interaction with sex (separate analyses for women and men can be found in **Supplementary Figure 3**).

With regard to consumption of different alcoholic beverages (Figure 4b and 5b), beer was statistically significantly associated with 4 pathways (the superpathway of sulfur oxidation, L-glutamate degradation via hydroxyglutarate, ppGpp biosynthesis and L-lysine biosynthesis), wine with 3 bacteria (*L. asaccharolyticus*, *S. mutans* and *B. dentium*) and 2 pathways (L-lysine biosynthesis and succinate fermentation to butanoate), and spirits with 1 bacterium (*F. bacterium GAG 95*) and 1 pathway (L-glutamate and L-glutamine biosynthesis). For the other beverage types, no statistical differences were detected.

### The gut microbiome as a potential mediator

Based on output from differential abundance analysis, an alcohol-associated microbial score was developed to examine the potential mediating role of alcohol-related bacteria in colorectal carcinogenesis (see **Supplementary Tables 6-7** for bacterial species significantly associated with alcohol intake overall and using the previously described five-fold cross-validation approach). The causal mediation analysis showed that the alcohol-associated microbial score partially mediated the association between alcohol intake and advanced colorectal lesions (**Figure 6a**). The total effect (95% CI) of alcohol on advanced lesions was 0.033 (0.017, 0.047), the average direct effect (ADE) was 0.029 (0.012, 0.043) and the average causal mediation effect (ACME) was 0.004 (0.001, 0.008). Overall, the proportion mediated was 12.1% (3.1, 32.2; **Figure 6b**), confirmed using an approach based on nested counterfactuals (here the proportion mediated was 12.2%, **Supplementary Table 8**). To assess potential differences in mediation effects between women and men while maintaining power, we conducted a mediation analysis omitting sex as a covariate. This did, however, only marginally affect the results.

**Figure 6.**
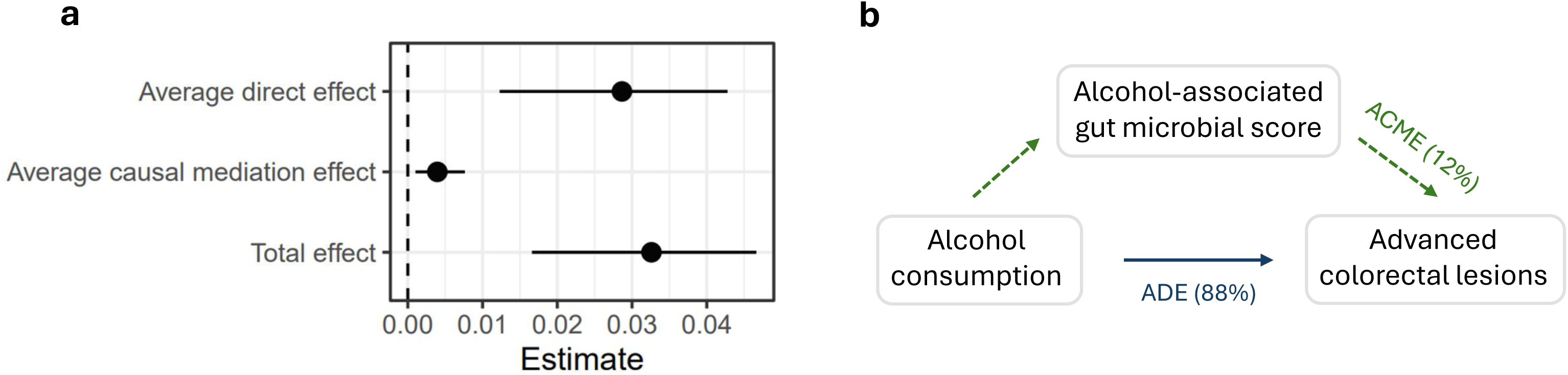
Causal mediation analysis.

## Discussion

This study provides new evidence on the detrimental role of alcohol in the development of colorectal lesions. We found a positive association between alcohol consumption and advanced colorectal lesions detected at screening even at modest intake levels. This association was strong in the overall population and in women, while no clear association was observed in men.

Consumers of alcohol had a distinct gut microbial profile, characterized by higher species diversity, altered microbial composition and differentially abundant bacteria and pathways. This distinctive microbial profile partly explained the association between alcohol intake and advanced lesions observed. Our results support a role of the microbiome in alcohol-induced colorectal carcinogenesis.

In the present study, every 10 g per day increase in alcohol consumption was associated with a 9% increased probability of advanced lesions being detected at colonoscopy, consistent across lesion subtype and location. Our results are in line with the literature on precancerous colorectal lesions. Meta-analyses have demonstrated a 27% increased risk of adenoma per 25 g alcohol consumed per day^36^, along with comparable increases in the risk of serrated polyps^37,38^.

Together, these results coincide with the literature on alcohol and CRC^29,39,40^.

In our study, consumption levels even below 10 g/day were associated with advanced lesions. As such, our findings reinforce cancer prevention guidelines of complete abstinence to achieve the lowest possible risk^41^.

The stronger association observed in women compared to men contrasts with prior literature. In the CUP 2016 meta-analysis from WCRF/AICR^29^, as well as two pooled analyses (from UK and Japan, respectively)^42,43^, no particular heterogeneity was found between sexes. Women from

Europe and Australia have the highest per-capita alcohol consumption worldwide, surpassing the global average by a factor of about two^5^. It is noteworthy that these regions also bear the highest incidence rates of CRC among women, Norway ranking at the top in 2020^44^. The consumption pattern of participants included in the present study seems to mirror those of the general Norwegian population^45,46^. Whether country-specific trends in alcohol consumption may account for divergence in findings remains a matter of speculation.

Our study demonstrated notable differences in the gut microbiome of participants consuming relative to not consuming alcohol, potentially of relevance to tumorigenesis. A link between alcohol intake and gut microbiome perturbations has been documented by others^47–51^. However, this has typically been studied in the context of chronic alcoholism^47,48,50^ and/or presence of severe liver pathologies^48,49^ that could confound the alcohol-gut microbiome relationship. We observed that even modest alcohol consumption was associated with an increase in α- and a convergence of β-diversity. The diversity change has also been reported in studies of other Western populations^52–55^, revealing alcohol as a strong source of gut microbiome variation.

We identified four bacterial species to be independently associated with alcohol consumption: *L. asaccharolyticus* and *B. finegoldii* were positively associated, whereas *C. symbiosum* and *S. mutans* were negatively associated. These results reproduce associations reported in the PREDICT study, which assessed microbial relationships with habitual diet in a large population, also finding *L. asaccharolyticus* and *C. symbiosum* to be strong determinants of alcohol consumption^55^. Among the identified bacteria, *C. symbiosum* has earlier been suggested as a potential biomarker for CRC^56,57^.

Interestingly, we found the association between alcohol consumption and detection of advanced lesions to be partially mediated by the gut microbiome. The mediated proportion was a modest, but conservative, 12% of the total effect, but nonetheless suggests that microbial changes caused by alcohol consumption could contribute to CRC development. How the relatively small concentrations of ethanol reaching the large intestine can induce microbial changes of relevance to carcinogenic development is however unclear. A conventional belief has been that the intestinal bacteria play an important role in metabolizing the remaining amounts of ethanol, through bacterial catalases and ADH activity, leading to accumulation of acetaldehyde and thus causing local damage^7,8,58^. However, this was recently questioned by Martino, *et al.*^6^ who used a mouse model to demonstrate that rather than metabolizing ethanol directly, gut bacteria responded to ethanol-feeding by activating acetate dissimilation. In line with these findings, we observed that TCA cycle gene abundance was positively associated with alcohol consumption. Acetate has recently received renewed attention in the context of CRC^7^. Although historically being regarded as protective, recent evidence suggests that acetate may contribute to cancer cell growth by serving as a substrate for the synthesis of acetyl-CoA^7^. We also found alcohol consumption to be associated with increased abundance of sulfur oxidation superpathway genes, regardless of consumption levels. Sulfur metabolism is characteristic of a wide range of bacteria^59^ and has been associated with CRC^59^ and other gut disorders^60^. In combination, these results suggest a potential for alcohol consumption to increase risk of CRC via bacterial acetate and sulfur metabolism.

A major strength of our study includes its large set of microbiome samples obtained through state-of-the-art methodology, coupled with validated exposure information. Access to clinically verified outcome data facilitated thorough investigations of screening-relevant outcomes, with minimal risk of misclassification. With a study population solely consisting of FIT positive participants, the proportion of detected lesions was high (63%). Nonetheless, this selective inclusion of participants may have restricted the generalizability of our findings. Other limitations must also be considered. First, the cross-sectional design limits causal interpretations, and the results must be viewed as hypothesis-generating only. Thus, although access to comprehensive data on lifestyle and demography allowed for detailed covariate adjustment (being particularly unique for the microbiome analyses), we cannot exclude the possibility of residual or reverse confounding. Second, selective inclusion of participants with colon bleeding at specimen collection, could have introduced bias. The proportion of participants with gastrointestinal morbidity may have been unevenly distributed between alcohol intake and outcome categories. This may be of particular concern for the present study, as some over- representation of former drinkers and “sick quitters” among the non-consumers, is to be expected^61^. However, excluding participants with self-reported gastrointestinal morbidity did not alter the observed associations.

To conclude, our study confirms the role of alcohol in the etiology of CRC. Consistent and positive associations were observed between alcohol consumption and advanced lesions even at moderate consumption levels, and particularly in women. Consuming alcohol was associated with a distinct microbial profile in the gut, manifested as increased species diversity, altered microbial composition and differentially abundant bacteria and pathways. Collectively, alcohol-associated bacteria mediated 12% of the association between alcohol intake and advanced colorectal lesions. The potential role of alcohol-associated microbial alterations in cancer development should be further examined in prospective cohort studies with long-term follow-up. Such studies should investigate potential sex differences and ideally expand the repertoire of biological mechanisms by evaluating metabolic, inflammatory, and immune-modulatory pathways.

## Data Availability

DNA sequencing data analyzed in this study are deposited in the database Federated EGA under accession code EGAS50000000170 (https://ega-archive.org/studies/EGAS50000000170). Per participant consent, submitted FASTQ files exclude reads mapping to the human genome. The data are available under restricted access due to the sensitive nature of data derived from human subjects. Processing of data from this study must comply with the General Data Protection Regulation (GDPR). Access can be obtained by following the procedure described here: https://www.mn.uio.no/sbi/english/groups/roungegroup/crcbiome/. Requests for data access can also be directed to Trine B Rounge.

## Acknowledgements

We would like to thank the devoted healthcare personnel, technicians and administrative staff at the two screening centers (Moss center and Bærum center) and university hospitals (Oslo University Hospital and Akershus University Hospital) for their important contributions to the BSCN and CRCbiome study. We offer special thanks to Jan Inge Nordby and Vahid Bemanian for their instrumental role in sample preparation and laboratory work. Library preparation and sequencing were carried out at the FIMM Technology Centre supported by HiLIFE and Biocenter Finland. We would particularly express our gratitude to Harri A. Kangas and Pekka J. Ellonen for their good cooperation, service and support. All processing of dietary data, including food and nutrient calculations, was conducted at the Department of Nutrition, University of Oslo. We would particularly like to thank Anne Marte Wetting Johansen for her dedicated work and attention to detail. Lastly, this article is the outcome of collective teamwork and scientific discussions among both former and current colleagues within our research group. Therefore, we would like to acknowledge Elina Vinberg, Even Sannes Riiser, Erik Natvig, Paula Istvan and Maja Jacobsen for their valuable contributions.

## Authors’ contributions

ASK, EB, EB, CBJ, TBR and PB designed the research (project conception, development of overall research plan, and study oversight).

ASK, EB, EA, CBJ, KRR, TBR and PB conducted the research (hands-on conduct of the experiments and data collection).

ASK, EB, CBJ, AH, KRR, GH, TBR and PB provided essential materials (applies to authors who contributed by providing animals, constructs, databases, etc, necessary for the research).

ASK, EB, CBJ and EB analyzed data or performed statistical analysis. ASK wrote the paper (only authors who made a major contribution). ASK and PB had the primary responsibility for the final content.

ASK, EB, EA, EB, CBJ, MDK, AH, CMP, JRH, MS, KRR, GH, TNR and PB have read and approved the final manuscript.

## Declaration of generative AI and AI-assisted technologies in the writing process

During the preparation of this work the first author(s) used Chat-GTP in order to improve language and readability. After using this tool/service, the author(s) reviewed and edited the content as needed and take(s) full responsibility for the content of the publication.

## Abbreviations

ACME: Average causal mediation effect
ADE: Average direct effect
ADH: Alcohol dehydrogenase
AICR: American Institute for Cancer Research
ALDH: Acetaldehyde dehydrogenase
BCSN: Bowel Cancer Screening in Norway
BMI: Body mass index
CRC: Colorectal cancer
CUP: Continuous Update Project
CYP2E1: Cytochrome P450 2E1 pathway
E: Exposure
E%: Energy percentage
FFQ: Food frequency questionnaire
FIT: Fecal immunochemical test
KBS: Kostberegningssystem
MaAsLin: Microbiome multivariable associations with linear models
M: Mediator
O: Outcome
PCoA: Principal coordinate analysis
PERMANOVA: Permutational multivariate analysis of variance
Q: Quartile
TE: Total effect
TSS: Total sum scaling
WCRF: World Cancer Research Fund

