## Supplementary Material for "The association between alcohol consumption and colorectal carcinogenesis is partially mediated by the gut microbiome"

**Supplemental material**

**Summary of supplementary information**

**Supplemental Table 1.** Odds ratios (ORs) and 95% confidence intervals (CIs) for presence of precursor lesions by subtype (left panel) and location (right panel) relative to controls by level of alcohol consumption (n=1,486).

**Supplemental table 2.** Odds ratios (ORs) and 95% confidence intervals (CIs) for presence of non-advanced adenoma^1^ and advanced lesions^2^ relative to controls by level of alcohol consumption in energy percentage (E%) in the study population as a whole (n=1,486) and by sex (660 women, 826 men)^3^.

**Supplemental table 3.** Odds ratios (ORs) and 95% confidence intervals (CIs) for presence of non-advanced adenoma^1^ and advanced lesions^2^ relative to controls by level of alcohol consumption in the study population as a whole (n=1,486) and by sex (660 women, 826 men)^3^. Missing values are imputed using multivariate imputation.

**Supplemental table 4.** Odds ratios (ORs) and 95% confidence intervals (CIs) for presence of non-advanced adenoma^1^ and advanced lesions^2^ relative to controls by level of alcohol consumption in the subset of participants with microbiome samples (n=947)^3^.

**Supplemental table 5.** Diversity and composition of the gut microbiome by level of alcohol consumption overall (n=947)^1^.

**Supplemental table 6.** Species association^1^ with alcohol intake in univariate and multivariate analysis.

**Supplemental table 7.** Species association^1^ with alcohol intake using five-fold cross-validation^2^ for generation of an alcohol-associated microbial score.

**Supplemental table 8.** Results of causal mediation analysis using the R package medflex.

**Supplemental Figure 1.** Covariation between the different alcohol consumption variables.

**Supplemental Figure 2.** Overlap between bacterial species and pathways linked to alcohol consumption and presence of advanced lesions.

**Supplemental Figure 3**. Differential abundance analyses of bacterial species and pathways by alcohol intake in women and men.

| **Supplemental Table 1.** Odds ratios (ORs) and 95% confidence intervals (CIs) for presence of precursor lesions by subtype (left panel) and location (right panel) relative to controls by level of alcohol consumption (n=1,486). | | | | | | | | | |
| --- | --- | --- | --- | --- | --- | --- | --- | --- | --- |
|  | **Control**  **(n=548)** | **Any advanced adenoma**  **(n=275)^1^** | | **Any advanced serrated lesion**  **(n=111)^2^** | | **Any proximal advanced lesion**  **(n=150)^3^** | | **Any distal advanced lesion**  **(n=241)^4^** | |
|  | n | n | OR (95% CI) | n | OR (95% CI) | n | OR (95% CI) | n | OR (95% CI) |
| Level of intake |  |  |  |  |  |  |  |  |  |
| 0 g/day | 88 | 22 | Ref. | 7 | Ref. | 13 | Ref. | 16 | Ref. |
| >0-10 g/day | 226 | 103 | **1.75 (1.02, 2.98)** | 46 | **2.47 (1.06, 5.80)** | 55 | 1.57 (0.80, 3.06) | 93 | **2.20 (1.21, 4.00)** |
| ≥10-20 g/day | 126 | 76 | **2.22 (1.25, 3.91)** | 29 | **2.81 (1.14, 6.92)** | 39 | 1.95 (0.96, 3.99) | 67 | **2.76 (1.46, 5.19)** |
| ≥20 g/day | 108 | 74 | **2.17 (1.21, 3.89)** | 29 | **2.84 (1.13, 7.10)** | 43 | **2.08 (1.01, 4.30)** | 65 | **2.75 (1.44, 5.24)** |
| *p_trend_* |  |  | **0.012** |  | 0.07 |  | **0.038** |  | **0.006** |
| Per 10 g increase/day |  |  | 1.09 (1.00, 1.20) |  | 1.05 (0.92, 1.20) |  | 1.06 (0.95, 1.19) |  | 1.10 (1.00. 1.21) |
| *Pcont* |  |  | 0.06 |  | 0.45 |  | 0.31 |  | 0.05 |
| Per 2-fold increase/day |  |  | **1.16 (1.05, 1.27)** |  | 1.13 (0.99, 1.29) |  | **1.12 (1.00, 1.26)** |  | **1.18 (1.07, 1.30)** |
| *Pcont* |  |  | **0.002** |  | 0.06 |  | **0.044** |  | **0.001** |

*Odds ratios (ORs) and 95% confidence intervals (CIs) are obtained from multinomial logistic regression analyses including the following clinical groups:*

*^1^Control (n=548), non-advanced adenoma (n=524), advanced serrated lesion only (n=74),* ***any advanced adenoma (n=275)*** *and CRC (n=65).*

*^2^Control (n=548), non-advanced adenoma (n=524), advanced adenoma only (n=238),* ***any advanced serrated lesion (n=111)*** *and CRC (n=65).*

*^3^Control (n=548), non-advanced adenoma (n=524), advanced distal lesion only (n=199****), any proximal advanced lesion (n=150)*** *and CRC (n=65).*

*^4^Control (n=548), non-advanced adenoma (n=524), advanced proximal lesion only (n=108),* ***any distal advanced lesion (n=241)*** *and CRC (n=65).*

*Complete adjustment set: age (continuous), sex, national affiliation (Norwegian affiliation, non-Norwegian affiliation, missing), screening center (center 1, center 2), education level (primary school, high school, college/university, missing), family history of CRC (yes, no, unknown/missing), smoking status (non-smoker, smoker, missing), BMI (continuous with missing set to median) and level of physical activity (continuous with missing set to median).*

| **Supplemental table 2.** Odds ratios (ORs) and 95% confidence intervals (CIs) for presence of non-advanced adenoma^1^ and advanced lesions^2^ relative to controls by level of alcohol consumption in energy percentage (E%) in the study population as a whole (n=1,486) and by sex (660 women, 826 men)^3^. | | | | | |
| --- | --- | --- | --- | --- | --- |
|  | **Control**  **(n=548)** | **Non-advanced adenoma**  **(n=524)** | | **Advanced lesions**  **(n=414)** | |
|  | n | n | OR (95% CI) | n | OR (95% CI) |
| **Overall** |  |  |  |  |  |
| Level of intake |  |  |  |  |  |
| 0 E% | 88 | 65 | Ref. | 34 | Ref. |
| >0-3 E% | 213 | 196 | 1.18 (0.80, 1.75) | 161 | **1.86 (1.18, 2.94)** |
| ≥3-6 E% | 128 | 117 | 1.14 (0.74, 1.75) | 109 | **2.04 (1.24, 3.33)** |
| ≥6 E% | 119 | 146 | 1.40 (0.91, 2.14) | 110 | **1.99 (1.21, 3.26)** |
| *p_trend_* |  |  | 0.14 |  | **0.039** |
| Per 3E% increase |  |  | 1.05 (0.97, 1.14) |  | **1.09 (1.00, 1.18)** |
| *Pcont* |  |  | 0.24 |  | **0.047** |
| Per 2-fold increase/day |  |  | 1.04 (0.97, 1.12) |  | **1.12 (1.04, 1.22)** |
| *Pcont* |  |  | 0.27 |  | **0.004** |
| **Men** |  |  |  |  |  |
| Level of intake |  |  |  |  |  |
| 0 E% | 31 | 22 | Ref. | 17 | Ref. |
| >0-3 E% | 89 | 113 | 1.60 (0.85, 3.02) | 87 | 1.61 (0.82, 3.18) |
| ≥3-6 E% | 68 | 66 | 1.16 (0.59, 2.26) | 76 | 1.84 (0.91, 3.72) |
| ≥6 E% | 79 | 104 | 1.47 (0.77, 2.80) | 74 | 1.44 (0.72, 2.89) |
| *p_trend_* |  |  | 0.74 |  | 0.65 |
| Per 3E% increase |  |  | 1.03 (0.93, 1.13) |  | 1.04 (0.94, 1.15) |
| *Pcont* |  |  | 0.59 |  | 0.44 |
| Per 2-fold increase/day |  |  | 1.00 (0.90, 1.11) |  | 1.07 (0.96, 1.20) |
| *Pcont* |  |  | 1.00 |  | 0.21 |
| **Women** |  |  |  |  |  |
| Level of intake |  |  |  |  |  |
| 0 E% | 57 | 43 | Ref. | 17 | Ref. |
| >0-3 E% | 124 | 83 | 0.97 (0.58, 1.62) | 74 | **2.11 (1.11, 3.98)** |
| ≥3-6 E% | 60 | 51 | 1.31 (0.73, 2.34) | 33 | **2.06 (1.00, 4.25)** |
| ≥6 E% | 40 | 42 | 1.53 (0.83, 2.83) | 36 | **3.15 (1.52, 6.55)** |
| *p_trend_* |  |  | 0.08 |  | **0.008** |
| Per 3E% increase |  |  | 1.09 (0.94, 1.25) |  | **1.20 (1.04, 1.39)** |
| *Pcont* |  |  | 0.26 |  | **0.012** |
| Per 2-fold increase/day |  |  | 1.10 (0.98, 1.23) |  | **1.20 (1.06, 1.36)** |
| *Pcont* |  |  | 0.10 |  | **0.003** |

*^1^Includes any adenoma (adenomatous polyp) not fulfilling the criteria of being advanced.*

*^2^Includes advanced adenoma, defined as any adenoma with either villous histology (≥25% villous components), high-grade dysplasia or polyp size greater than or equal to 10 mm; advanced serrated lesions, defined as any serrated lesions with size ≥ 10 mm or dysplasia; and colorectal cancer, defined as presence of adenocarcinoma arising from the colon or rectum.*

*^3^Odds ratios (ORs) and 95% confidence intervals (CIs) are obtained from multinomial logistic regression analyses adjusting for the following covariates: age (continuous), sex (except in the sex-specific analyses), national affiliation (Norwegian affiliation, non-Norwegian affiliation, missing), screening center (center 1, center 2), education level (primary school, high school, college/university, missing), family history of CRC (yes, no, unknown/missing), smoking status (non-smoker, smoker, missing), BMI (continuous with missing set to median) and level of physical activity (continuous with missing set to median).*

| **Supplemental table 3.** Odds ratios (ORs) and 95% confidence intervals (CIs) for presence of non-advanced adenoma^1^ and advanced lesions^2^ relative to controls by level of alcohol consumption in the study population as a whole (n=1,486) and by sex (660 women, 826 men)^3^. Missing values are imputed using multivariate imputation. | | | | | |
| --- | --- | --- | --- | --- | --- |
|  | **Control**  **(n=548)** | **Non-advanced adenoma**  **(n=524)** | | **Advanced lesions**  **(n=414)** | |
|  | n | n | OR (95% CI) | n | OR (95% CI) |
| **Overall** |  |  |  |  |  |
| Level of intake |  |  |  |  |  |
| 0 g/day | 88 | 65 | Ref. | 34 | Ref. |
| >0-10 g/day | 226 | 202 | 1.16 (0.79, 1.71) | 164 | **1.82 (1.15, 2.86)** |
| ≥10-20 g/day | 126 | 131 | 1.31 (0.86, 1.99) | 104 | **1.98 (1.21, 3.24)** |
| ≥20 g/day | 108 | 126 | 1.30 (0.84, 2.02) | 112 | **2.18 (1.32, 3.61)** |
| *p_trend_* |  |  | 0.19 |  | **0.010** |
| Per 10 g increase/day |  |  | 1.03 (0.94, 1.12) |  | **1.09 (1.00, 1.18)** |
| *Pcont* |  |  | 0.53 |  | **0.049** |
| Per 2-fold increase/day |  |  | 1.03 (0.96, 1.11) |  | **1.13 (1.05, 1.23)** |
| *Pcont* |  |  | 0.438 |  | **0.002** |
| **Men** |  |  |  |  |  |
| Level of intake |  |  |  |  |  |
| 0 g/day | 31 | 22 | Ref. | 17 | Ref. |
| >0-10 g/day | 87 | 111 | 1.58 (0.84, 2.96) | 83 | 1.58 (0.80, 3.11) |
| ≥10-20 g/day | 65 | 73 | 1.40 (0.72, 2.70) | 74 | 1.94 (0.96, 3.91) |
| ≥20 g/day | 84 | 99 | 1.36 (0.71, 2.58) | 80 | 1.50 (0.75, 2.99) |
| *p_trend_* |  |  | 0.90 |  | 0.47 |
| Per 10 g increase/day |  |  | 1.01 (0.92, 1.11) |  | 1.05 (0.95, 1.15) |
| *Pcont* |  |  | 0.85 |  | 0.35 |
| Per 2-fold increase/day |  |  | 0.99 (0.90, 1.10) |  | 1.09 (0.98, 1.21) |
| *Pcont* |  |  | 0.902 |  | 0.116 |
| **Women** |  |  |  |  |  |
| Level of intake |  |  |  |  |  |
| 0 g/day | 57 | 43 | Ref. | 17 | Ref. |
| >0-10 g/day | 139 | 91 | 0.93 (0.57, 1.53) | 81 | **2.09 (1.12, 3.91)** |
| ≥10-20 g/day | 61 | 58 | 1.39 (0.79, 2.45) | 30 | 1.78 (0.86, 3.67) |
| ≥20 g/day | 24 | 27 | 1.58 (0.78, 3.20) | 32 | **4.65 (2.11, 10.24)** |
| *p_trend_* |  |  | 0.07 |  | **0.001** |
| Per 10 g increase/day |  |  | 1.08 (0.90, 1.29) |  | **1.26 (1.05, 1.50)** |
| *Pcont* |  |  | 0.41 |  | **0.013** |
| Per 2-fold increase/day |  |  | 1.08 (0.96, 1.21) |  | **1.21 (1.07, 1.37)** |
| *Pcont* |  |  | 0.182 |  | **0.003** |

*^1^Includes any adenoma (adenomatous polyp) not fulfilling the criteria of being advanced.*

*^2^Includes advanced adenoma, defined as any adenoma with either villous histology (≥25% villous components), high-grade dysplasia or polyp size greater than or equal to 10 mm; advanced serrated lesions, defined as any serrated lesions with size ≥ 10 mm or dysplasia; and colorectal cancer, defined as presence of adenocarcinoma arising from the colon or rectum.*

*^3^Odds ratios (ORs) and 95% confidence intervals (CIs) are obtained from multinomial logistic regression analyses adjusting for the following covariates: age (continuous), sex (except in the sex-specific analyses), national affiliation (Norwegian affiliation, non-Norwegian affiliation), screening center (center 1, center 2), education level (primary school, high school, college/university), family history of CRC (yes, no), smoking status (non-smoker, smoker), BMI (continuous) and level of physical activity (continuous). For covariates with missing data (national affiliation, education level, family history of CRC, smoking status, BMI and level of physical activity), values were imputed using the R package MICE (Multivariate Imputation via Chained Equations), assuming that the missing data were missing at random.*

| **Supplemental table 4.** Odds ratios (ORs) and 95% confidence intervals (CIs) for presence of non-advanced adenoma^1^ and advanced lesions^2^ relative to controls by level of alcohol consumption in the subset of participants with microbiome samples (n=947)^3^. | | | | | |
| --- | --- | --- | --- | --- | --- |
|  | **Control**  **(n=414)** | **Non-advanced adenoma**  **(n=130)** | | **Advanced lesions**  **(n=403)** | |
|  | n | n | OR (95% CI) | n | OR (95% CI) |
| **Overall** |  |  |  |  |  |
| Level of intake |  |  |  |  |  |
| 0 g/day | 67 | 19 | Ref. | 32 | Ref. |
| >0-10 g/day | 181 | 47 | 0.85 (0.45, 1.60) | 160 | **1.76 (1.07, 2.87)** |
| ≥10-20 g/day | 95 | 30 | 0.89 (0.44, 1.80) | 102 | **1.93 (1.13, 3.30)** |
| ≥20 g/day | 71 | 34 | 1.33 (0.65, 2.71) | 109 | **2.72 (1.57, 4.72)** |
| *p_trend_* |  |  | 0.26 |  | **0.001** |
| Per 10 g increase/day |  |  | 1.09 (0.95, 1.25) |  | **1.14 (1.03, 1.27)** |
| *Pcont* |  |  | 0.23 |  | **0.010** |
| Per 2-fold increase/day |  |  | 1.03 (0.91, 1.16) |  | **1.17 (1.07, 1.28)** |
| *Pcont* |  |  | 0.69 |  | **<0.001** |

*^1^Includes any adenoma (adenomatous polyp) not fulfilling the criteria of being advanced.*

*^2^Includes advanced adenoma, defined as any adenoma with either villous histology (≥25% villous components), high-grade dysplasia or polyp size greater than or equal to 10 mm; advanced serrated lesions, defined as any serrated lesions with size ≥ 10 mm or dysplasia; and colorectal cancer, defined as presence of adenocarcinoma arising from the colon or rectum.*

*^3^Odds ratios (ORs) and 95% confidence intervals (CIs) are obtained from multinomial logistic regression analyses adjusting for the following covariates: age (continuous), sex (except in the sex-specific analyses), national affiliation (Norwegian affiliation, non-Norwegian affiliation, missing), screening center (center 1, center 2), education level (primary school, high school, college/university, missing), family history of CRC (yes, no, unknown/missing), smoking status (non-smoker, smoker, missing), BMI (continuous with missing set to median) and level of physical activity (continuous with missing set to median).*

.

| **Supplemental table 5.** Diversity and composition of the gut microbiome by level of alcohol consumption overall (n=947)^1^. | | | | | | | |
| --- | --- | --- | --- | --- | --- | --- | --- |
|  | **α-diversity** | | | | **β-diversity** | | |
|  | Shannon | | Inverse Simpson | | Bray-Curtis dissimilarity | | |
|  | Median (Q1, Q3) | β (95% CI) | Median (Q1, Q3) | β (95% CI) | R^2^ | Ω^2^ | p-value |
| **Overall alcohol intake** |  |  |  |  |  |  |  |
| Consumers/non-consumers |  |  |  |  | **0.00206** | **0.00103** | **0.004** |
| Non-consumers | 3.1 (2.9, 3.3) | 0 (reference) | 13.1 (9.5, 17.1) | 0 (reference) |  |  |  |
| Consumers | 3.2 (3.0, 3.4) | **2.7 (0.6, 4.8)** | 14.6 (11.3, 18.5) | **10.2 (1.5, 19.7)** |  |  |  |
| Increasing level of intake |  |  |  |  | **0.00405** | **0.00093** | **0.030** |
| 0 g/day | 3.1 (2.9, 3.3) | 0 (reference) | 13.1 (9.5, 17.1) | 0 (reference) |  |  |  |
| >0-10 g/day | 3.2 (3.0, 3.4) | **2.8 (0.5, 5.0)** | 14.7 (11.0, 18.4) | **11.1 (1.9, 21.2)** |  |  |  |
| ≥10-20 g/day | 3.2 (3.0, 3.4) | 2.3 (-0.2, 4.7) | 14.2 (11.5, 18.1) | 8.7 (-1.2, 19.6) |  |  |  |
| ≥20 g/day | 3.3 (3.0, 3.4) | **3.0 (0.5, 5.5)** | 15.0 (11.2, 18.8) | 9.7 (-0.5, 20.9) |  |  |  |
| *p_trend_* |  | 0.123 |  | 0.312 |  |  |  |
| Per 10 g increase/day | - | 0.1 (-0.3, 0.5) | - | 0.0 (-1.6, 1.7) | **0.00157** | **0.00053** | **0.044** |
| *Pcont* | - | 0.628 | - | 0.983 |  |  |  |
| Per 2-fold increase/day | - | 0.3 (-0.1, 0.7) | - | 0.7 (-0.9, 2.4) | **0.00193** | **0.00089** | **0.002** |
| *Pcont* | - | 0.117 | - | 0.380 |  |  |  |
| Adherence to guidelines |  |  |  |  | **0.00310** | **0.00102** | **0.014** |
| Fully adhering (0 g/day) | 3.1 (2.9, 3.3) | 0 (reference) | 13.1 (9.5, 17.1) | 0 (reference) |  |  |  |
| Partially adhering (<10/20 g/day) | 3.2 (3.0, 3.4) | **2.7 (0.6, 4.9)** | 14.5 (11.1, 18.3) | **10.6 (1.6, 20.4)** |  |  |  |
| Not adhering (≥10/20g /day) | 3.2 (3.0, 3.4) | **2.6 (0.3, 5.0)** | 14.8 (11.3, 18.6) | 9.5 (-0.1, 19.9) |  |  |  |
| *p_trend_* |  | 0.106 |  | 0.172 |  |  |  |
| **Alcohol subtypes** |  |  |  |  |  |  |  |
| **Beer** |  |  |  |  |  |  |  |
| Consumers/non-consumers |  |  |  |  | 0.00141 | 0.00037 | 0.081 |
| Non-consumers | 3.2 (2.9, 3.4) | 0 (reference) | 13.9 (10.1, 17.8) | 0 (reference) |  |  |  |
| Consumers | 3.2 (3.0, 3.4) | **2.3 (0.7, 3.8)** | 14.6 (11.5, 18.5) | **7.2 (1.0, 13.9)** |  |  |  |
| Per unit increase/day | - | 0.6 (-0.4, 1.6) | - | 1.9 (-2.0, 5.9) | 0.00137 | 0.00033 | 0.119 |
| *Pcont* | - | 0.242 | - | 0.341 |  |  |  |
| Per 2-fold increase/day | - | **0.3 (0.1, 0.6)** | - | **1.0 (0.1, 1.9)** | **0.00162** | **0.00058** | **0.029** |
| *Pcont* | - | **0.003** | - | **0.031** |  |  |  |
| **Wine** |  |  |  |  |  |  |  |
| Consumers/non-consumers |  |  |  |  | 0.00143 | 0.00039 | 0.084 |
| Non-consumers | 3.2 (3.0, 3.4) | 0 (reference) | 14.0 (10.4, 17.8) | 0 (reference) |  |  |  |
| Consumers | 3.2 (3.0, 3.4) | 1.2 (-0.4, 2.8) | 14.6 (11.4, 18.3) | 4.4 (-1.9, 11.1) |  |  |  |
| Per unit increase/day | - | -0.3 (-1.3, 0.7) | - | -1.8 (-5.7, 2.3) | **0.00151** | **0.00047** | **0.045** |
| *Pcont* | - | 0.527 | - | 0.384 |  |  |  |
| Per 2-fold increase/day | - | 0.1 (-0.1, 0.4) | - | 0.4 (-0.6, 1.3) | **0.00186** | **0.00083** | **0.002** |
| *Pcont* | - | 0.371 | - | 0.456 |  |  |  |
| **Spirits** |  |  |  |  |  |  |  |
| Consumers/non-consumers |  |  |  |  | 0.00094 | -0.00011 | 0.607 |
| Non-consumers | 3.2 (3.0, 3.4) | 0 (reference) | 14.2 (10.5, 18.0) | 0 (reference) |  |  |  |
| Consumers | 3.2 (3.1, 3.4) | **1.9 (0.4, 3.5)** | 14.9 (12.0, 18.7) | **6.7 (0.5, 13.2)** |  |  |  |
| Per unit increase/day | - | 0.8 (-1.0, 2.7) | - | -0.4 (-7.4, 7.2) | 0.00127 | 0.00023 | 0.206 |
| *Pcont* | - | 0.378 | - | 0.920 |  |  |  |
| Per 2-fold increase/day | - | 0.4 (-0.1, 0.8) | - | 0.7 (-1.1, 2.5) | 0.00092 | -0.00013 | 0.676 |
| *Pcont* | - | 0.117 | - | 0.448 |  |  |  |
| **Drinks** |  |  |  |  |  |  |  |
| Consumers/non-consumers |  |  |  |  | 0.00124 | 0.00020 | 0.199 |
| Non-consumers | 3.2 (3.0, 3.4) | 0 (reference) | 14.5 (10.8, 18.3) | 0 (reference) |  |  |  |
| Consumers | 3.2 (3.0, 3.4) | 0.2 (-1.5, 2.0) | 14.2 (11.7, 18.0) | -0.3 (-7.0, 7.0) |  |  |  |
| Per unit increase/day | - | 0.5 (-2.6, 3.6) | - | 4.1 (-7.9, 17.7) | 0.00099 | -0.00006 | 0.534 |
| *Pcont* | - | 0.758 | - | 0.518 |  |  |  |
| Per 2-fold increase/day | - | 0.0 (-0.3, 0.4) | - | 0.0 (-1.4 (1.4) | 0.00125 | 0.00021 | 0.188 |
| *Pcont* | - | 0.845 | - | 0.960 |  |  |  |
| **Non-alcoholic drinks** |  |  |  |  |  |  |  |
| Consumers/non-consumers |  |  |  |  | 0.000867 | -0.00018 | 0.738 |
| Non-consumers | 3.2 (3.0, 3.4) | 0 (reference) | 14.3 (10.8, 18.1) | 0 (reference) |  |  |  |
| Consumers | 3.2 (3.1, 3.4) | 1.5 (-0.1, 3.1) | 14.7 (11.7, 18.7) | 4.5 (-1.8, 11.1) |  |  |  |
| Per unit increase/day | - | -0.3 (-4.6, 4.3) | - | -3.4 (-19.1, 15.4) | 0.00117 | 0.00013 | 0.315 |
| *Pcont* | - | 0.908 | - | 0.704 |  |  |  |
| Per 2-fold increase/day | - | 0.2 (-0.1, 0.5) | - | 0.5 (-0.6, 1.6) | 0.00086 | -0.00019 | 0.738 |
| *Pcont* | - | 0.182 | - | 0.404 |  |  |  |

*^1^Regression coefficients (β’s) and 95% confidence intervals (CIs) are obtained from multivariate linear regression analyses adjusting for the following covariates: age (continuous), sex, national affiliation (Norwegian affiliation, non-Norwegian affiliation, missing), screening center (center 1, center 2), education level (primary school, high school, college/university, missing), family history of CRC (yes, no, unknown/missing), smoking status (non-smoker, smoker, missing), BMI (continuous with missing set to median), level of physical activity (continuous with missing set to median) and sequencing depth (continuous). The diversity indices were log-transformed prior to analyses to improve normalization and ease interpretation. Eta-squared (R^2^), omega-squared (Ω*^2^*) and p-values were derived from permutational multivariate analyses of variance (PERMANOVA) based on the Bray-Curtis dissimilarity index with adjustment for the same set of covariates as listed above. Relevant packages and functions included vegan::adonis2 and micEco::adonis_OmegaSq.*

*.*

| **Supplemental table 6.** Species association^1^ with alcohol intake in univariate and multivariate analysis. Only species significantly associated with alcohol consumption (either positively or negatively) are included in the table. | | |
| --- | --- | --- |
| **Species** | Univariate | Multivariate |
| *Agathobaculum butyriciproducens* | **+** |  |
| *Alistipes putredinis* | **+** |  |
| *Alistipes shahii* | **+** |  |
| *Bacteroides finegoldii* | **+** | **+** |
| *Bifidobacterium dentium* | **-** | **-** |
| *Blautia sp CAG 257* | **-** |  |
| *Butyricimonas virosa* | **+** |  |
| *Clostridium innocuum* | **-** |  |
| *Clostridium symbiosum* | **-** | **-** |
| *Coprobacter fastidiosus* | **+** |  |
| *Coprococcus comes* | **+** |  |
| *Eggerthella lenta* | **-** |  |
| *Eisenbergiella tayi* | **-** |  |
| *Enterocloster bolteae (Clostridium bolteae)* | **-** | **-** |
| *Enterocloster citroniae (Clostridium citroniae)* | **-** |  |
| *Enterocloster clostridioformis (Clostridium clostridioformis)* | **-** |  |
| *Erysipelatoclostridium ramosum* | **-** |  |
| *Faecalibacterium prausnitzii* | **+** |  |
| *Lawsonibacter asaccharolyticus* | **+** | **+** |
| *Paraprevotella xylaniphila* | **+** |  |
| *Phocaeicola dorei* | **+** |  |
| *Phocaeicola vulgatus (Bacteroides vulgatus)* | **+** |  |
| *Roseburia sp CAG 182* | **+** |  |
| *Ruminococcus gnavus* | **-** |  |
| *Sellimonas intestinalis* | **-** |  |
| *Streptococcus anginosus group* | **-** |  |
| *Streptococcus mutans* | **-** | **-** |
| *Streptococcus oralis* | **-** |  |
| *Streptococcus vestibularis* | **-** |  |

*^1^Results are derived from differential abundance analyses using microbiome multivariable associations with linear models (MaAsLin) 2. The complete adjustment set included the following covariates: age (continuous), sex, national affiliation (Norwegian affiliation, non-Norwegian affiliation, missing), screening center (center 1, center 2), education level (primary school, high school, college/university, missing), family history of CRC (yes, no, unknown/missing), smoking status (non-smoker, smoker, missing), BMI (continuous with missing set to median) and level of physical activity (continuous with missing set to median). ^2^All participants were included in the data set for this analysis.*

| **Supplemental table 7.** Species association^1^ with alcohol intake using five-fold cross-validation^2^ for generation of an alcohol-associated microbial score. | | | | | | | |
| --- | --- | --- | --- | --- | --- | --- | --- |
| **Species** | CV 1 | CV 2 | CV 3 | CV 4 | CV 5 | | Summary |
| *Asaccharobacter celatus* |  |  |  |  | **+** | | **1/5** |
| *Bacteroides finegoldii* | **+** | **+** |  |  |  | | **2/5** |
| *Bacteroides massiliensis* |  | **+** |  |  |  | | **1/5** |
| *Bifidobacterium dentium* | **-** | **-** | **-** | **-** |  | | **4/5** |
| *Clostridium innocuum* |  |  | **-** |  |  | | **1/5** |
| *Clostridium saccharolyticum* | **+** |  |  |  |  | | **1/5** |
| *Clostridium spiroforme* |  |  |  |  | **+** | | **1/5** |
| *Clostridium symbiosum* |  | **-** | **-** | **-** |  | | **3/5** |
| *Eggerthella lenta* |  |  | **-** |  |  | | **1/5** |
| *Eisenbergiella tayi* | |  | **-** | **-** |  |  | **2/5** |
| *Enterocloster bolteae (Clostridium bolteae)* | |  | **-** | **-** | **-** | **-** | **4/5** |
| *Enterocloster clostridioformis (Clostridium clostridioformis)* | |  | **-** |  |  |  | **1/5** |
| *Erysipelatoclostridium ramosum* |  |  | **-** |  |  | | **1/5** |
| *Eubacterium sp_CAG_38* |  |  | **+** |  |  | | **1/5** |
| *Finegoldia magna* |  |  |  | **-** |  | | **1/5** |
| *Lawsonibacter asaccharolyticus* | **+** | **+** | **+** | **+** | **+** | | **5/5** |
| *Streptococcus mutans* | **-** |  | **-** | **-** | **-** | | **4/5** |

*^1^Results are derived from differential abundance analyses using microbiome multivariable associations with linear models (MaAsLin) 2. The complete adjustment set included the following covariates: age (continuous), sex, national affiliation (Norwegian affiliation, non-Norwegian affiliation, missing), screening center (center 1, center 2), education level (primary school, high school, college/university, missing), family history of CRC (yes, no, unknown/missing), smoking status (non-smoker, smoker, missing), BMI (continuous with missing set to median) and level of physical activity (continuous with missing set to median). ^2^Data were assigned to five non-overlapping partitions, where differential abundance was assessed for four of five partitions, and results are reported for each held-out partition. An alcohol-associated microbial score was calculated for the held-out partition based on the abundance of the indicated species.*

| **Supplemental table 8.** Results of causal mediation analysis using the R package medflex. Results are presented as natural direct effect, natural indirect effect, total effect and propotion mediated. | | | |
| --- | --- | --- | --- |
|  | Estimate (95% CI) | Z value | p-value |
| Natural direct effect | 1.14 (1.05-1.24) | 3.11 | 0.00185 |
| Natural indirect effect | 1.02 (1.00, 1.03) | 2.26 | 0.0241 |
| Total effect | 1.16 (1.07, 1.26) | - | - |
| Proportion mediated | 0.122 |  |  |

*Alcohol intake (per 2-fold increase/day) was regarded as the primary exposure, the alcohol associated gut microbial score as the mediator, and advanced colorectal lesions as the outcome. The analyses were adjusted for age (continuous), sex, national affiliation (Norwegian affiliation, non-Norwegian affiliation, missing), screening center (center 1, center 2), education level (primary school, high school, college/university, missing), family history of CRC (yes, no, unknown), smoking status (non-smoker, smoker, missing), BMI (continuous with missing set to median) and level of physical activity (continuous with missing set to median).*

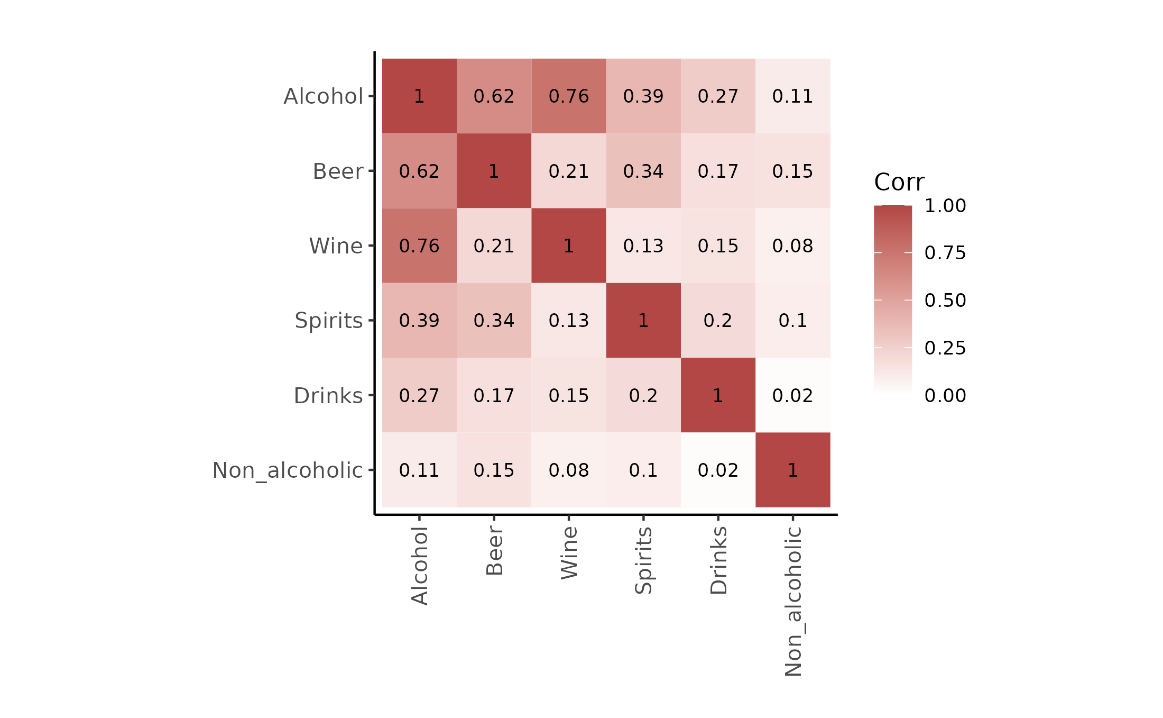

**Supplemental Figure 1.** Covariation between the different alcohol consumption variables. Values are Spearman’s correlation coefficients.

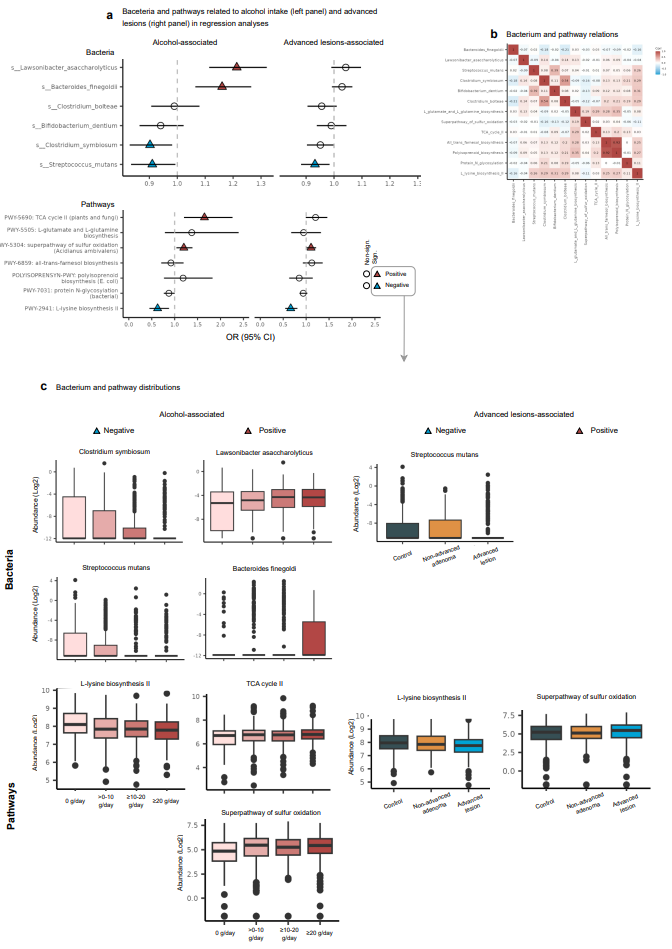

**Supplemental Figure 2.** Overlap between bacterial species and pathways linked to alcohol consumption and presence of advanced lesions. a) Bacterial species and pathways identified as significant predictors of alcohol consumption in differential abundance analyses were examined for their independent associations with alcohol (left panel) and advanced lesions (right panel) in multinomial logistic regression models with adjustment for the following covariates: age (continuous), sex, national affiliation (Norwegian affiliation, non-Norwegian affiliation, missing), screening center (center 1, center 2), education level (primary school, high school, college/university, missing), family history of CRC (yes, no, unknown/missing), smoking status (non-smoker, smoker, missing), BMI (continuous with missing set to median), level of physical activity (continuous with missing set to median) and the other candidate species and pathways (as continuous variables). Effect estimates represent odds ratios with 95% confidence intervals. Significant results (p-value <0.05) are marked with a triangle formation, with color indicating the direction of the association (red: positive, blue: negative). B) Correlation matrix of bacterial species and pathways identified as significant predictors of alcohol consumption. Values are Spearman’s correlation coefficients. c) Distribution of bacterial species and pathways identified as significant predictors of alcohol consumption (left) or advanced lesions (right) in the regression analyses described under point a) by consumption level (left) and colonoscopy finding (right). Values were log2-transformed with a pseudo-count of half the minimum value prior to visualization.

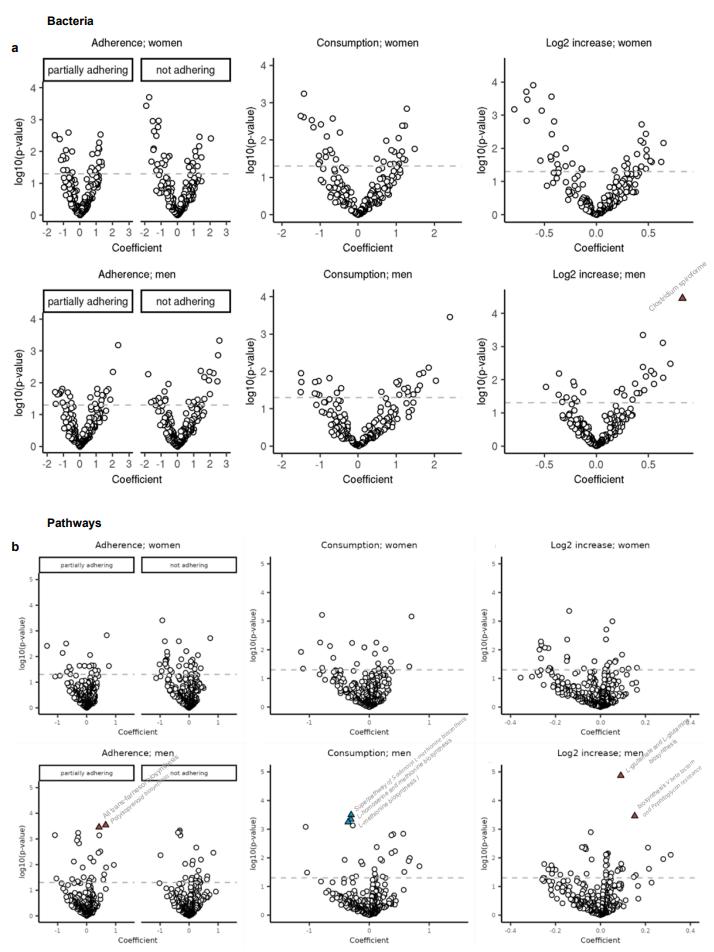

**Supplemental Figure 3**. Differential abundance analyses of bacterial species and pathways by alcohol intake in women and men. Volcano plots showing the multivariate adjusted associations for bacterial species (a) and pathways (b) by the following three measures of alcohol intake: adherence to alcohol guidelines (with “Full adherence” as reference category), consumption relative to non-consumption (with “non-consumption” as reference category) and per 2-fold increase in intakes. Associations were studied using microbiome multivariable associations with linear models (MaAsLin) 2. Values on the x-axis represent log2 fold changes, while values on the y-axis represent log10-transformed p-values. The dotted line is positioned at -log10(0.05), indicating significance (non-corrected p-values). Species and pathways significant also after Benjamini-Hochberg (BH) correction are marked with a triangular formation, with color indicating the direction of the association (red: positive, blue: negative).
